# Effects of parental autoimmune diseases on type 1 diabetes in offspring can be partially explained by HLA and non-HLA polymorphisms

**DOI:** 10.1101/2024.04.16.24305884

**Authors:** Feiyi Wang, Aoxing Liu, Zhiyu Yang, Pekka Vartiainen, Sakari Jukarainen, Satu Koskela, Richard A. Oram, Lowri Allen, Jarmo Ritari, Jukka Partanen, Markus Perola FinnGen, Tiinamaija Tuomi, Andrea Ganna

## Abstract

Type 1 diabetes (T1D) and other autoimmune diseases (AIDs) often co-occur in families. Leveraging data from 58,284 family trios in Finnish nationwide registers (FinRegistry), we identified that, out of 50 parental AIDs examined, 15 were associated with an increased T1D risk in offspring. These identified epidemiological associations were further assessed in 470K genotyped Finns from FinnGen study through comprehensive genetic analyses, partitioned into HLA and non-HLA variations. Using FinnGen’s 12,563 trios, a within-family polygenic transmission analysis demonstrated that the aggregation of many parental AIDs with offspring T1D can be partially explained by HLA and non-HLA polymorphisms in a disease-dependent manner. We, therefore, proposed a parental polygenic score (PGS), incorporating both HLA and non-HLA polymorphisms, to characterize the cumulative risk pattern of T1D in offspring. This raises an intriguing possibility of using parental PGS, in conjunction with clinical diagnoses, to inform individuals about T1D risk in their offspring.

## INTRODUCTION

Type 1 diabetes (T1D) is an autoimmune disease (AID) characterized by a severe insulin deficiency resulting from the destruction of insulin-producing pancreatic beta cells. While its pathogenicity remains largely unknown, T1D usually appears in genetically susceptible individuals and is triggered by environmental exposures^1–3^ - mostly onset before the age of 20, with incidence increasing from birth and peaking at age 10 to 14 during puberty.^4–6^ Notably, in many populations such as Europeans, Asians, and Latins, genetic polymorphisms in human leukocyte antigen (HLA) genes were reported to account for up to half of the T1D heritability, for which the strongest associations were attributable to two HLA class II haplotypes - *DR3-DQ2* and *DR4-DQ8*.^7–11^ In addition to the well-documented significant HLA contribution, non-HLA polymorphisms also play a role in T1D inheritance^12^ – with large-scale genome-wide association studies (GWAS), dozens of independent non-HLA signals have been identified across the genome.^12–14^ This, together with HLA associations, can be used to construct polygenic scores (PGS) to inform individuals’ genetic predisposition of T1D prior to the disease onset^15–17^ as well as better differentiate T1D from other types of diabetes.^18–20^

Interestingly, T1D often co-occur with other AIDs, both in the same individuals and within families (Supplementary notes 1). Previous studies have explored this co-occurrence from two main approaches. A popular and straightforward one adopts a population-based design and uses epidemiological analyses to study the risk of T1D among people whose relatives had AIDs. We have provided a detailed review of such studies as Table S1. To obtain conclusive findings, a sufficient number of trios or families is needed. ^21–23^ Another approach attempts to leverage genetic information for a better understanding of T1D-AID aggregation among genetically related individuals.^24–26^ For example, the *HLA-DR3-DQ2* haplotype was more likely to be seen among T1D children having celiac disease (CD) relatives in their extended family.^25^ Whereas most of the studies mainly collected family history from questionnaires and focused only on HLA haplotypes, others systematically researched the disease associations of common variants in either HLA or non-HLA genes. Notably, rather than individual-level data, many studies utilized publicly accessible summary statistics generated from population-based analyses. For example, a recent study discovered significant intercorrelations among several AIDs including T1D, rheumatoid arthritis (RA), and autoimmune hypothyroidism (HYPO), in two large HLA pleiotropy hotspots.^27^ Further, GWAS revealed shared genetic susceptibility loci beyond the HLA regions for T1D and several other AIDs such as CD, RA, and HYPO.^28–30^

To date, there has been a lack of comprehensive investigation that combines the two approaches and encompasses a wider range of AID spectrum other than CD, RA, and HYPO. Also, it remains an enigma why some individuals with presumably similar genetic backgrounds can develop such heterogeneous diseases ranging from potentially lethal insulin deficiency to mild hypothyroidism - even among members of the same family who also tend to share similar environmental factors. One way to approach this question is to combine evidence from population-based multi-generational cohorts and large-scale biobanks with high-resolution diagnostic data, preferably of the same population, to avoid any potential bias or confounding relevant to differences in across-population socio-cultural contexts and clinical practices. In this study, we aim to comprehensively evaluate the genetic determinants of familial aggregation of T1D and other AIDs using the multi-generational health registers of the whole Finnish population in FinRegistry,^31^ as well as the genomic and family trio data available in Finnish biobanks through the FinnGen study.^32^ Considering the world’s highest incidence of T1D in Finland,^33^ these detailed and structured data resources provide us a unique opportunity to quantify the associations between parental AIDs and offspring T1D and, more importantly, to seek for answers of three key but underexplored questions: 1) Which parental diagnoses of AIDs are associated with T1D in offspring? 2) To what extent is the shared genetic background between AIDs and T1D driven by genetic components, separately for HLA and non-HLA regions? And 3) can AID-related genetic information of a person be used to estimate the T1D risk in his or her offspring?

## RESULTS

We leveraged nationwide socio-demographic and health registers of the entire Finnish population collected in the FinRegistry study^31^ (N = 7.2 million) and a subset of 473,681 genotyped Finns from the FinnGen study^32^ (Figure 1A and B). T1D and AIDs were identified based on a clinical diagnosis recorded in the health registers. We defined T1D cases as those who had their first diagnosis recorded before the age of 40 years; the majority also had a specific diagnosis for T1D recorded later when the specific International Classification of Diseases (ICD) codes were introduced. In total, we included 14,571 T1D cases in FinRegistry (0.61%; Figure S2.2) and 3,668 in FinnGen (0.83%). To examine the association between AIDs in parents and T1D in offspring, as well as the extent to which genetic factors could contribute to the identified association, we constructed 2.4 million family trios in FinRegistry based on Finnish multi-generation registers and 12,563 genotype-inferred trios in FinnGen (STAR Methods). Of these trios, 14,571 in FinRegistry and 1,129 in FinnGen had children with T1D (T1D-trios). To have broad coverage of different AIDs, we defined 50 AIDs or autoimmune-related disorders based on Finnish versions of the ICD codes 8-10 (Supplementary notes 2.1). After excluding 13 AIDs with less than 50 parental cases in FinRegistry and 11 AIDs that were highly correlated with other AIDs, we included 26 AIDs for further analyses (Figure 2; Table S2.1). The number of parental cases ranged from 55 for autoimmune hemolytic anemia (AIHA) (corresponding to a prevalence of 0.05%) to 11,485 for HYPO (prevalence = 9.85%) (Table S2.1; Table S3.1.1).

**Figure 1:**
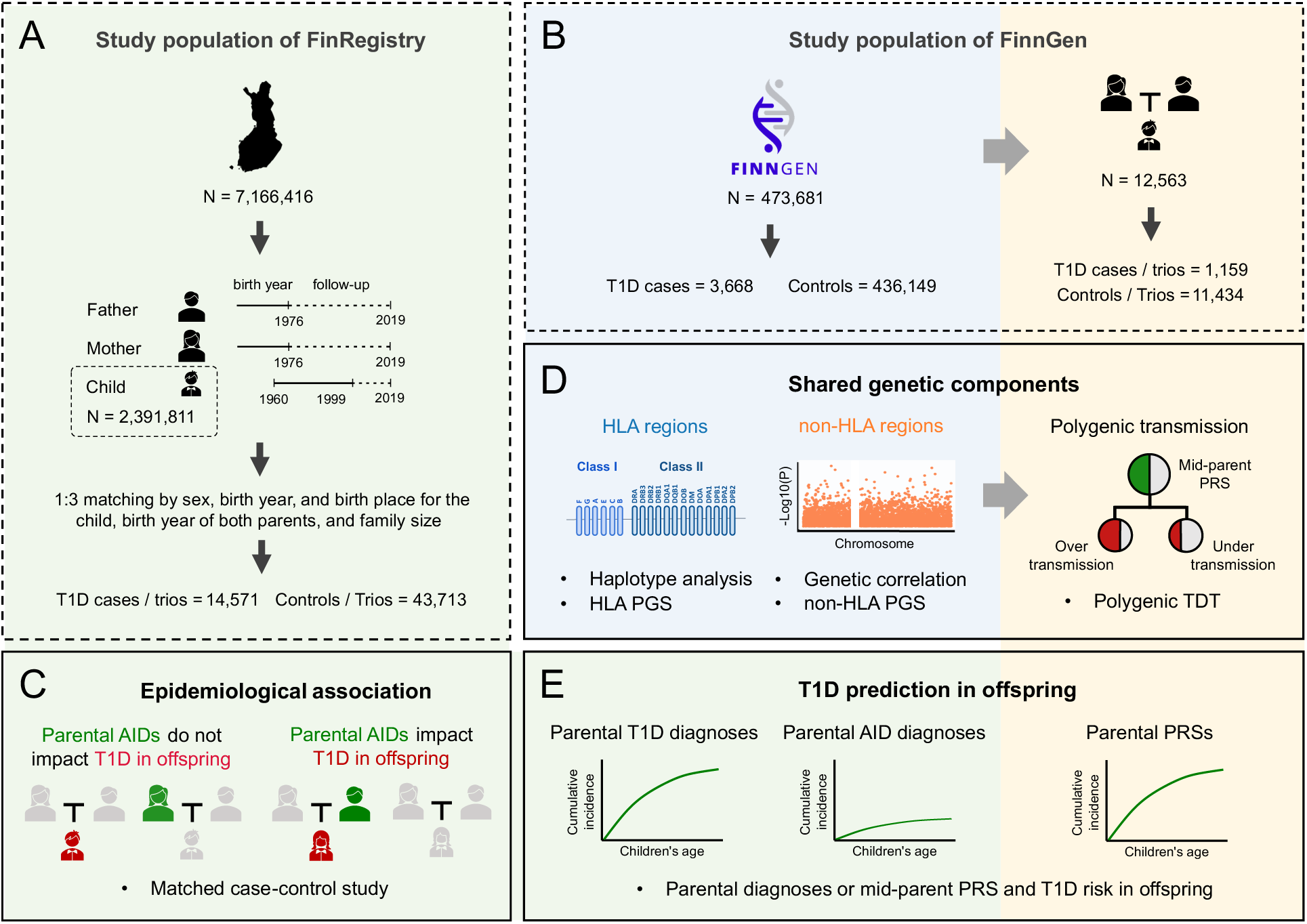
An overview of the study design and study populations. (A) The study population of FinRegistry (7.2 million individuals) represents every Finn alive in 2010 (5.3 million individuals) and their first-degree relatives. To maximize the coverage of diagnoses of AIDs for parents and T1D for children, we included only family trios with both parents born before 1976 and children between 1960-1999 (the follow-up time in 2019 was at least 45 years for parents and 20 years for children). The solid lines denote the birth year range of the study population, and the dashed lines are the years of follow-up. Among the 2.4 million family trios, 14,571 had a child ever diagnosed with T1D (T1D trio). For each T1D-trio, we matched three control trios based on sex, birth year, birthplace, and the number of siblings of the child, as well as the birth year of both parents. In total, we included 14,571 T1D-trios and 43,713 matched control trios. (B) FinnGen comprises 473.7K Finns enrolled through a nationwide network of biobanks. We studied 3,370 T1D cases, 385,786 controls, as well as 12,563 family trios constructed with genomic information (1,129 T1D-trios and 11,434 control trios). (C) A matched case-control study was conducted among the 58,284 FinRegistry family trios to examine the association between parents’ AID and offspring’s TID. The different symbols denote father, mother, and offspring, and color the disease status (red, offspring with T1D; green, parents with AIDs; grey, individuals unaffected by T1D or other AIDs). (D) Shared genetic components between T1D and other AIDs were examined using population-based analyses (on the left with a blue background) or trio-based family analyses (on the right with a yellow background). A haplotype and PGS-based analysis of HLA variants was conducted in 473.7K FinnGen participants (left); a genetic correlation analysis of the non-HLA variants utilized GWAS summary statistic data (middle); and a polygenic transmission disequilibrium test (pTDT) examined in FinnGen participants whether the AID-associated common variants as a whole were over-transmitted from AID-unaffected parents to their T1D-affected offspring (right). (E) The average AID PGS of the parents and its predictive performance of T1D in offspring among 12,563 genotyped family trios in FinnGen.

**Figure 2:**
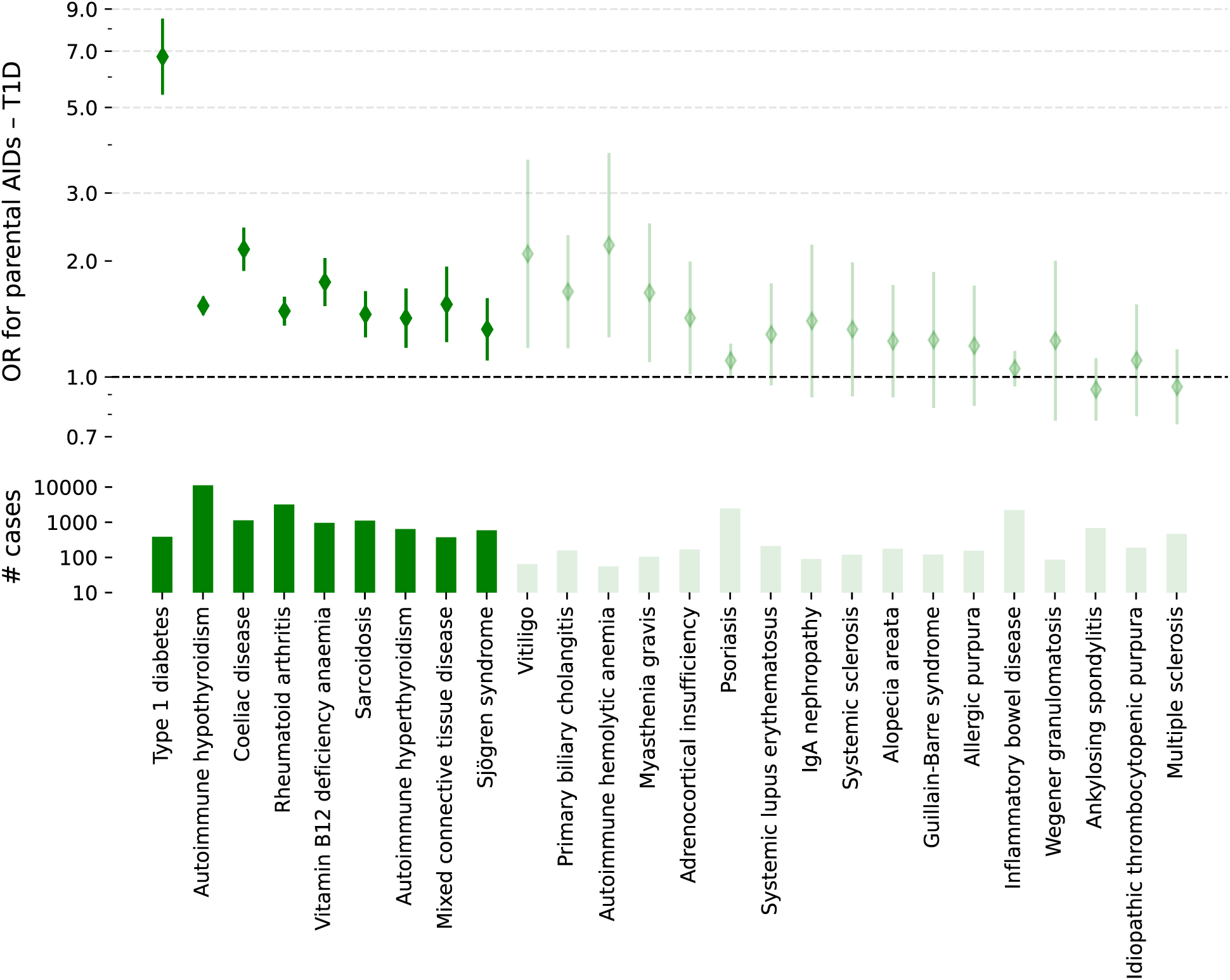
Epidemiological associations between parental AIDs and T1D in offspring ordered by P values. Upper panel: association (OR, and 95% CI) between parental AIDs and T1D in offspring using a matched case-control design in Finnish nationwide registry data (FinRegistry). Lower panel: number of T1D cases with parents having a given AID diagnosis. The dark green diamonds and bars indicate AIDs that are significantly associated with T1D in offspring after multiple testing corrections.

### Epidemiological associations between parental AIDs and T1D in offspring

We first examined the associations between parental AIDs and T1D in children using the family trios in the FinRegistry study. For each of the 14,571 trios with T1D-affected children, we employed a 1:3 matched case-control design considering the information of both the child (sex, birth year, birthplace, and number of siblings) and parents (birth year) (STAR Methods; Figure 1C). Regarding other socio-demographic factors that were not used in matching, we saw limited differences between the T1D trios and the control trios (Table S3.1.2). Overall, children with T1D (42.2% [95% CI 41.4%, 43.0%]) were more likely to have AID-affected parent(s) compared to those without T1D (31.9% [31.5%, 32.3%], P_difference_=9.9×10^--118^).

Of 26 parental AIDs examined in FinRegistry, 15 AIDs were associated with an increased risk of T1D in offspring at a nominal P value threshold (P<0.05), and nine remained statistically significant after Bonferroni correction for multiple testing (P = 0.05/26 = 0.002) (Figure 2; Table S3.2.1). The strongest association was seen for T1D (odds ratio (OR) [95% CI], 6.77 [5.44, 8.42], P=3.8×10^-66^), followed by CD (2.14 [1.90, 2.42], P=2.5×10^-35^) and vitamin B12 deficiency anemia (B12A, 1.76 [1.54, 2.02], P=1.4×10^-16^). In terms of sex, we didn’t observe significant differences either for parents or for offspring (Figure S3.2.2). However, consistent with previously studies,^34–36^ we observed that fathers with T1D were more likely than mothers with T1D to have children with T1D (71% vs 65%). Similarly, more children with T1D had fathers with T1D compared to mothers with T1D (1.2% vs 0.6%). A sensitivity analysis restricted to early-onset T1D (before age 20) yielded stronger associations for parental early-onset T1D and SLE than those in the full cohort (Table S3.2.4.1; Table S3.2.4.2).

### Shared genetic components from HLA and non-HLA variants at a population level

Parents and offspring share both genetic components and environmental exposures. We next aimed to quantify the extent to which the identified epidemiological associations are attributable to a shared genetic background. Given that T1D is known to be impacted by both HLA and non-HLA genes,^12–14^ we analyzed HLA and non-HLA variants separately at a population level.

#### Multi-allelic scores (PGS) of HLA alleles for AIDs and T1D

Having observed that multiple HLA haplotypes (Figure S3.3.1.3) and amino acids (Supplementary notes 3.3.2) could contribute to the susceptibility to T1D and other AIDs independently, we first constructed polygenic HLA scores (HLA PGSs) to summarize the overall multi-allelic HLA effects for each AIDs. Considering the widespread variation in HLA alleles across populations,^37^ we opted to construct HLA PGSs in FinnGen with 187 imputed Class I and II alleles using weighted ridge regression (STAR Methods). We were able to construct reliable HLA PGSs (P≤0.05/26 and partial correlation |ρ|≥2% for predicting the disease risk itself) in 23,336 individuals for 14 AIDs (including T1D). Of these, HLA PGSs for 13 AIDs exhibited significant associations with T1D susceptibility after multiple-test correction (0.05/14 = 0.003), with exceptions for psoriasis (P=3.6×10^-3^) and ankylosing spondylitis (P=6.1×10^-2^) (Figure 3A; Table S3.3.3.1; Table S3.3.3.2). Overall, the effects of AID HLA PGS on individuals’ own T1D risk were highly correlated with the effects of parental AIDs on offspring T1D obtained from our FinRegistry epidemiological analyses (ρ from a Spearman’s rank correlation = 0.63, P=6.1×10^-4^). Among all AIDs other than T1D (OR=5.33 [4.79, 5.93], P =3.74 × 10^-207^), the strongest associations were seen for adrenocortical insufficiency (OR=2.30 [2.13, 2.49], P =4.2×10^-100^), RA (OR=2.10 [1.95, 2.27], P=1.8×10^- 78^), and CD (OR=2.07 [1.94, 2.22], P =2.4×10^-96^). Consistent with the HLA haplotype analysis, we observed negative associations with T1D for HLA PGS of inflammatory bowel disease (IBD, OR=0.64 [0.60, 0.69], P=1.2 × 10^-30^) and multiple sclerosis (MS, OR=0.81 [0.75, 0.88], P=2.4 × 10^-7^). These associations were further replicated with a distinct analytic approach - leave-one-group-out (LOGO) (Figure S2.4.2; Table S3.3.3.3).

**Figure 3:**
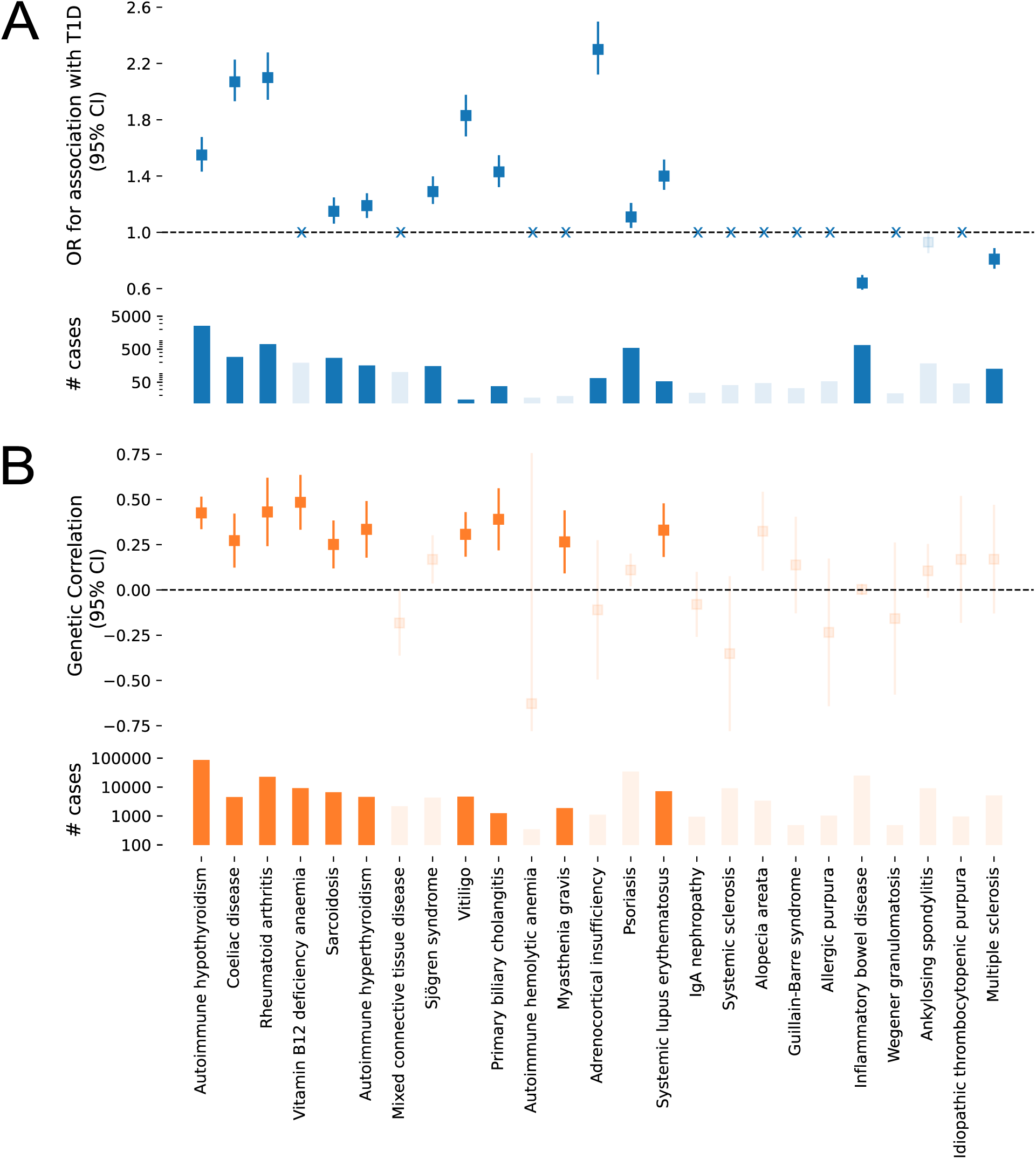
Shared genetic background between T1D and other AIDs, stratified by HLA and non-HLA variations at a population level. (A) Upper panel: association (OR, and 95% CI) between HLA PGSs for different AIDs and T1D in FinnGen. The dark blue squares indicate that HLA PGS for a given disease has a significant association with T1D after multiple testing corrections. “×” denotes that an HLA PGS could not be robustly constructed for that AID (STAR Methods). Bottom panel: blue bars represent the number of individuals with the given disease. (B) Upper panel: non-HLA based genetic correlations (r_g_ and 95% CI) between AIDs and T1D using GWAS summary statistics from European populations. Bottom panel: the number of cases for the given disease. The dark orange squares or bars indicate that the AIDs have a significant r_g_ with T1D after multiple testing corrections.

#### Genome-wide non-HLA correlation between T1D and AIDs

In addition to the major HLA contributors, non-HLA variants identified in the GWAS studies are known to impact AID susceptibility as well. We, therefore, explored the shared correlation between T1D and other AIDs using a summary statistics-based method - linkage disequilibrium score regression (LDSC),^38,39^ with the HLA regions excluded (STAR Methods). Significant positive genetic correlations (r_g_) were observed between T1D and ten AIDs (P<0.05/25 after Bonferroni correction) (Figure 3B), including HYPO, CD, RA, B12A, sarcoidosis, autoimmune hyperthyroidism (HYPER), vitiligo, primary biliary cholangitis (PBC), myasthenia gravis, and systemic lupus erythematosus (SLE). The first six AIDs were also among the parental AIDs that exerted significant effects on offspring T1D in our FinRegistry epidemiological analyses (ρ from a Spearman’s rank correlation=0.30, P=0.15). The highest r_g_ were seen for B12A (r_g_=0.48 [0.34, 0.63], P=6.4×10^-11^), followed by HYPO (0.43 [0.34, 0.51], P=2.9×10^-23^) and RA (0.43 [0.25, 0.61], P=3.9×10^-6^) (Table S3.3.4.1). Contrary to the negative HLA association, we did not observe a significant non-HLA genetic correlation between T1D and MS or between T1D and IBD. A sensitivity analysis excluding all chromosome 6 variants yielded similar estimates, suggesting that the observed non-HLA results were not driven by variants in highly linkage disequilibrium (LD) with HLA alleles (Table S3.3.4.2).

In summary, both the HLA and non-HLA analyses recapitulated the multi-generational epidemiological associations observed in nationwide registers. While the results conferred that the risk of T1D was positively associated with the risk of many other AIDs regarding both HLA and non-HLA genetic variants (e.g., HYPO, CD, RA), the opposite held for MS and IBD concerning the HLA susceptibility whereas no genetic correlation was observed in non-HLA regions.

### Transmitted and non-transmitted genetic liability within families

#### Polygenic transmission disequilibrium test for HLA and non-HLA genetic factors

Following the population-based epidemiological evidence for clustering of other AIDs and T1D in the trios, and the genetic evidence for a shared genetic origin of the AIDs and T1D at a population level, we wanted to examine the transmission of the HLA and non-HLA within families using a polygenic transmission disequilibrium test (pTDT).^40^ The pTDT examines whether the AID risk variants are observed to be over-transmitted (or under-transmitted) to the offspring with T1D (compared to the expected transmission rate), and thus, it is immune to many of the potential confounders arising from population studies on unrelated individuals. We considered 12,563 trios (genotyped in FinnGen), of which 1,159 had offspring with T1D (9.2%). The prevalence of T1D in these trios was higher than expected in the general population because FinnGen includes several studies that have specifically targeted individuals with T1D. We analyzed 10 AIDs with reliable PGSs (ρ≥2%) for both HLA and non-HLA genes and six additional diseases with only reliable PGSs for the HLA regions (STAR Methods; Table 3.3.3.2; Table S3.4.1). For offspring with T1D, both HLA and non-HLA PGSs for T1D deviated significantly from their mid-parent value (1.23 [1.16, 1.30], P=2.6×10^-168^ and 0.69 [0.62, 0.75], P=9.6×10^-74^) while no such deviation was observed in unaffected siblings (−0.06 [-0.13, 0.01], P=0.08 and 0.00 [-0.06, 0.05], P=0.88) (Figure 4; Table S3.4.2). Overall, two major patterns were seen across the analyzed AIDs. The first group encompassed T1D, HYPO, RA, and SLE, for which significant over-transmission in offspring with T1D was seen for both HLA and non-HLA PGSs. The second group exhibited significant transmission only for HLA but not non-HLA PGSs: while CD and psoriasis exhibited significant over-transmission in offspring with T1D compared to unaffected siblings, IBD and MS had a significant under-transmission. Taking IBD as an example, among 2,040 genotyped trios, 756 offspring with T1D presented under-transmitted HLA PGS (−0.38 [-0.45, -0.31], P=3.1×10^-22^) while their 1,269 unaffected siblings had comparable PGS to their parents (−0.01 [-0.08, 0.06], P=0.17). No diseases showed significant pTDT only regarding non-HLA PGS without a significant association for HLA-PGS.

**Figure 4:**
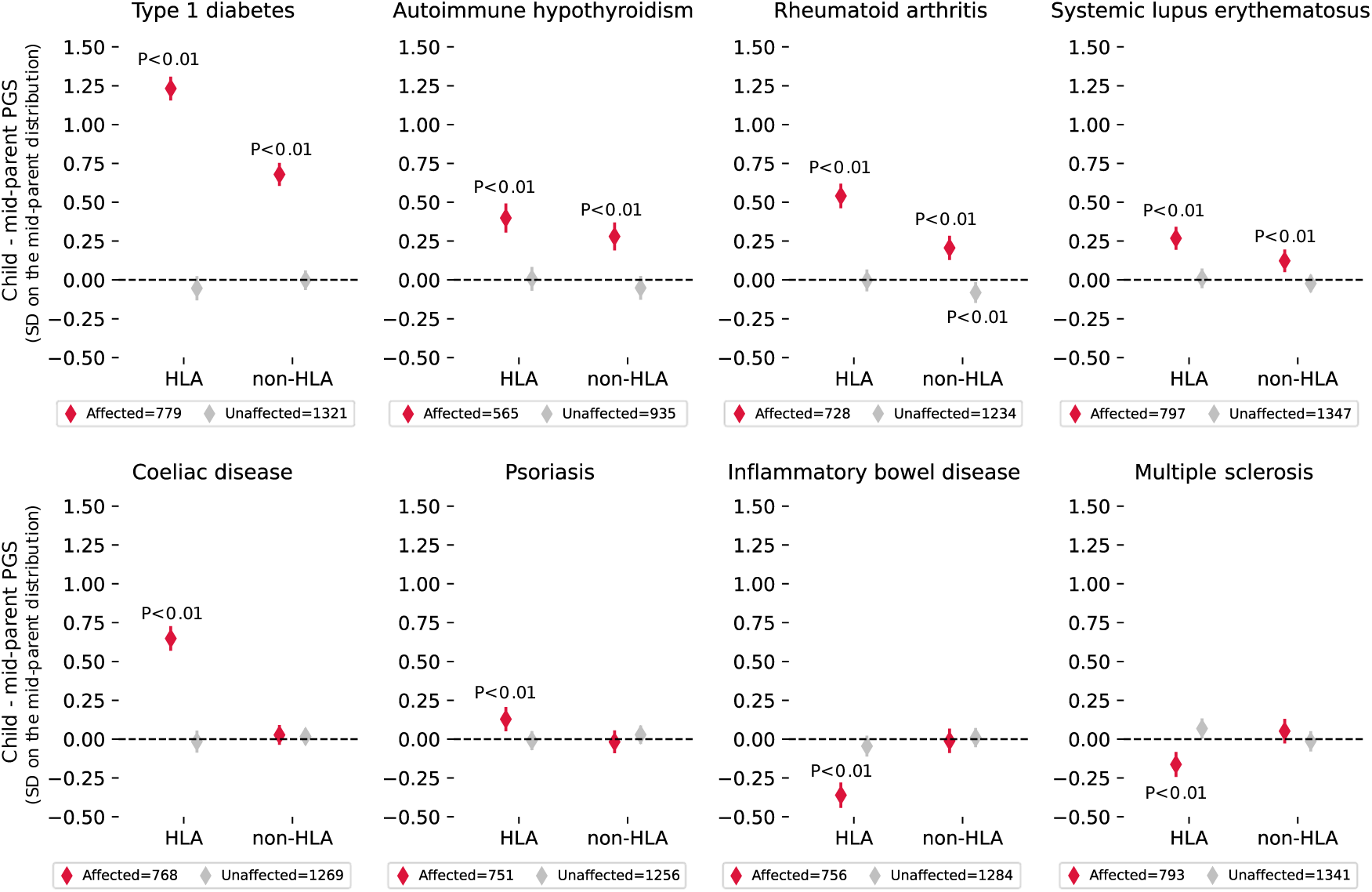
Polygenic transmission disequilibrium tests (pTDT) assess whether the transmission of AID PGSs to offspring significantly deviated from the mid-parent PGS, among T1D-affected offspring and their unaffected siblings. Deviations from mid-parent PGS (mean and 95% CI) of 1 SD change in offspring PGS are present, separately for HLA and non-HLA regions. The P value is obtained from a two-sided t-test and shown as “P<0.01” for significant results; otherwise, it is not marked. Red color denotes offspring with T1D and gray unaffected siblings.

### Offspring T1D risk based on clinical history and genetic information of parental AIDs

We asked whether a couple’s genetic information could be used to assess T1D risk in offspring. To address this question, we adopted a novel strategy to construct a full PGS (Full-PGS) by considering the contribution ratio of HLA PGS relative to non-HLA PGS (Supplementary notes 3.5). We tested eight AIDs (T1D, HYPO, RA, SLE, CD, psoriasis, IBD, and MS) for which either an HLA PGS or non-HLA PGS was reliable in predicting T1D (|ρ|≥1%) (Table S3.5.2) using a five-fold cross-validation design. By modeling the HLA and non-HLA variations separately and using imputed HLA alleles (187 alleles in 10 classical HLA genes rather than common genotyped SNPs), the Full-PGSs we proposed outperformed the standard SNP-based PGS methods such as PRS-CS^41^ when predicting the disease risk of individuals themselves (Table S3.6.1; Table S3.6.2) as well as when predicting T1D in offspring with AID-specific parental PGS among the 12,563 FinnGen genotyped trios (Table S3.6.3; Figure S3.6.4).

Having established that a parental Full-PGS for T1D was strongly associated with T1D risk in offspring (AUC_mean_=0.818; pseudo-R^2^_mean_=0.198), we tested whether the parental Full-PGSs for all the eight AIDs could add any additional information to the parental Full-PGS for T1D (AUC_mean_=0.820; pseudo-R^2^_mean_=0.202). While McFadden’s R^2^ shows a statistically significant difference between the two models (P_difference_=2.9 × 10^-26^), the small magnitude of this difference, along with the comparable AUCs (P_difference_=0.793), implied a limited improvement after integrating the PGS of eight AIDs. This suggested that, instead of a direct effect, the impact of parental AID PGS on offspring T1D was most likely mediated through the genetic correlation between T1D and AID and, therefore, provided limited extra information to the offspring T1D prediction when parental T1D PGS was already included in the model and sufficiently accurate.

We thus focused on parental Full-PGS for T1D and estimated the cumulative risk of T1D in offspring using a Cox proportional hazards model adjusted for the first 10 PCs and birth year of the child. When we used the mean birth year of all offspring (1984) and stratified PGS into four groups by percentiles (0-50^th^, 50-90^th^, 90-99^th^, 99-100^th^), we observed distinct trajectories in terms of T1D cumulative incidence rates (0.24%, 0.74%, 2.29%, 6.96% by age 20) (Figure 5A). That is, children whose parents were in the top percentile of T1D Full-PGS had a 29-fold higher risk of developing T1D than children whose parents were within the bottom 50% of T1D Full-PGS. When further stratified by the sex of children (Figure 5B and C), the adjusted survival curves suggested that sons overall had a higher cumulative incidence than daughters across all PGS groups. For example, in the top T1D parental Full-PGS group, sons (11.48%) could have a 2-fold higher cumulative risk than daughters (5.29%) by age 20. Overall, the cumulative incidence curves of T1D started to level off around the age of 14 to 16.

**Figure 5:**
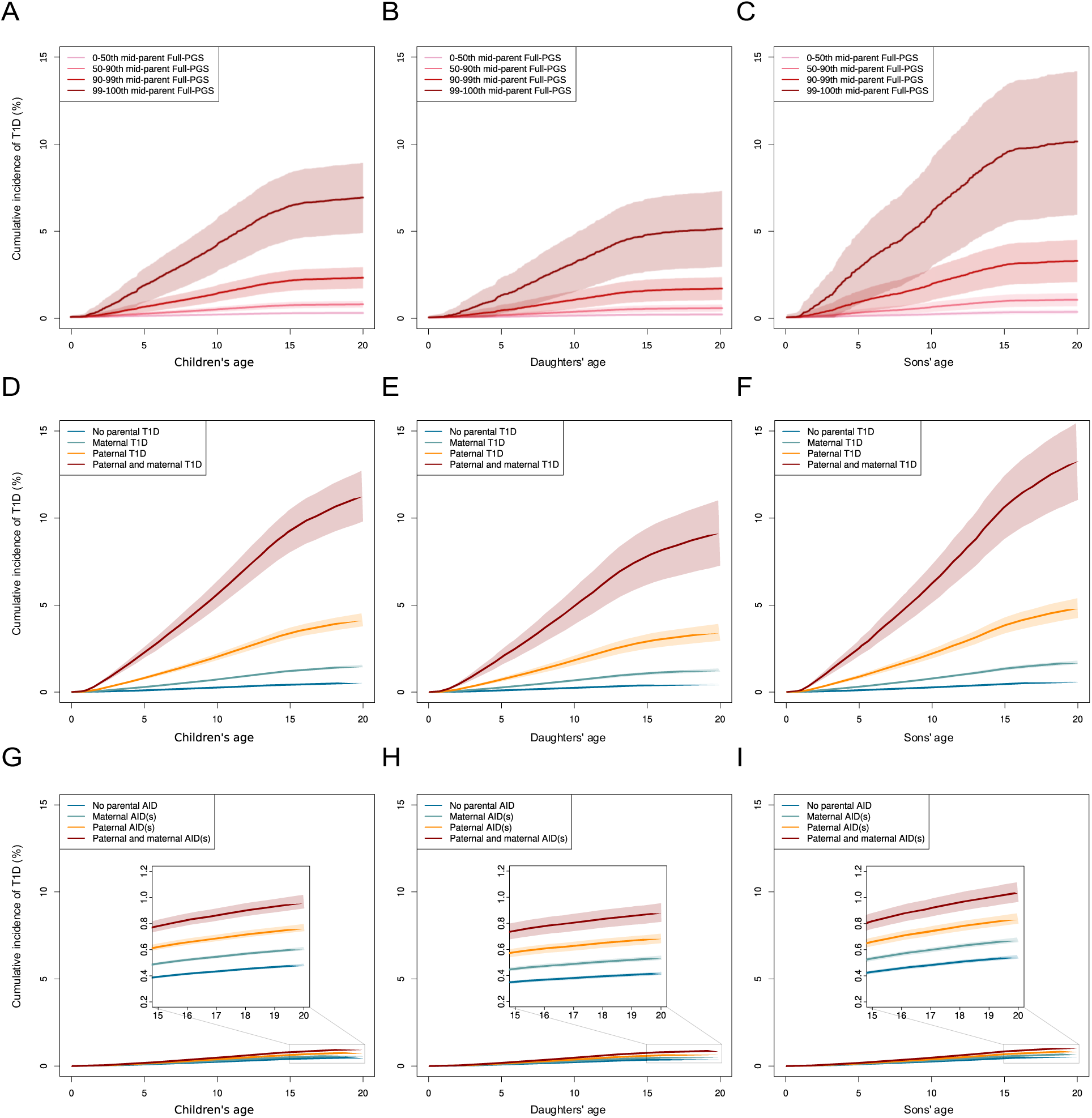
Adjusted survival curves with 95% confidence interval from Cox proportional hazards models for cumulative T1D incidence by age 20 in offspring, stratified by parental full T1D PGS percentiles comprising both HLA and non-HLA variants (**Panels A-C**; 8,872 FinnGen genotyped trios), parents’ T1D status (**Panels D-F;** 3,048,812 FinRegistry trios), parents’ other AIDs (**Panels G-I**; 3,037,723 FinRegistry trios after removing trios having parent(s) diagnosed with T1D), including T1D, autoimmune hypothyroidism, rheumatoid arthritis, systemic lupus erythematosus, coeliac disease, psoriasis, inflammatory bowel disease, and multiple sclerosis; The panels show pooled data for both sexes (left), daughters (middle), and sons (right).

We then wanted to understand how informative the Full-PGS was compared to parental AID clinical history. In general, the offspring of T1D couples had a higher cumulative incidence of T1D risk (Figure 5D) than T1D-unaffected couples and such differences were larger for boys (Figure 5E) than girls (Figure 5F). Having either father or mother with T1D (prevalence_father_ = 0.22%, prevalence_mother_ = 0.15%) resulted in a lower cumulative risk than having both parents in the top percentile of Full-PGS for T1D (1.49% and 4.16% vs 6.95% by age 20 for children). Having both parents affected by T1D (a very rare event with only a prevalence of 8.5 per million) resulted in a higher cumulative risk (11.30%) than having parents in the top percentile of Full-PGS for T1D (6.95%). Among couples only affected by AIDs other than T1D, offspring also had a higher cumulative incidence of T1D than the general population (Figure 5G, H, and I) (0.96 % vs 0.48%), albeit the cumulative incidence would be much smaller than that for the offspring of couples with diagnosed T1D or couples in the top decile of the Full-PGS for T1D. Thus, non-T1D AID diagnoses in parents offer limited additional value in assessing the T1D risk for their offspring.

## DISCUSSION

In this study, we comprehensively explored the genetic determinants of the familial aggregation of T1D and other AIDs with data from high-dimensional nationwide registers and rich genetic information of the Finnish population. Our long follow-up (≥20 years) in offspring and the early age of T1D onset (median age = 12.7) allows us to maximize the number of T1D cases identified in the study. With systematically designed epidemiological and genetic analyses, we seek to answer three key but underexplored questions: which parental AIDs are related to T1D in offspring, to what extent the identified parental AID-offspring T1D association is attributable to genetic polymorphisms of HLA and non-HLA variations, and last, for any couples that are planning to have children, how well we could use their genetic information to evaluate the T1D risk of their offspring.

The nationwide registers of 7.2 Finns collected in the FinRegistry study cover longitudinal health and sociodemographic information of 58,284 family trios, which provides us a unique opportunity to perform a comprehensive exploration of the impact of 50 autoimmune diseases on offspring T1D. The approach not only allowed a scale and detailedness inaccessible in questionnaire- or survey-based studies, but enabled an efficient matched case-control design to better control for unmeasurable confounding factors than the standard unmatched design used in previous large population-based studies.^21–23^ In total, we detected 10 parental AID-offspring T1D associations, including T1D, HYPO, CD, RA, HYPER, and B12A, that have previously been linked to T1D risk in offspring in large population-based studies (Table S4), and more importantly, six novel associations encompassing AIHA, mixed connective tissue disease, myasthenia gravis, alopecia areata, psoriasis, and vitiligo. Following the epidemiological analyses, we conducted a set of genetic analyses in a hypothesis-free manner to better understand the genetic causes of the observed family aggregations. We leveraged the genetic information of 470K genotyped Finns enrolled in the FinnGen study to study the shared effects of HLA and non-HLA variations at a population level and within families using pTDT in 12,563 FinnGen family trios, which, to the best of our knowledge, is the largest family-based genetic analysis performed for T1D and AIDs. Similar to the epidemiological family-based study that is widely accepted to yield less biased estimates than population-based studies, pTDT can condition out the impact of shared familial factors through within-family comparisons, allowing, for the first time, to examine under- or over-transmission of genetic risk factors for AIDs in individuals with T1D and their unaffected siblings.

Generally, two major patterns were seen regarding the transmission of AID-associated variants to offspring with T1D, with one group encompassing T1D, HYPO, RA, and SLE showing significant over-transmission for both HLA and non-HLA variants. The second group exerted significant deviated transmission only for HLA variants, with over-transmission in the case of CD and psoriasis, and under-transmission in the case of IBD and MS. Overall, the epidemiological disease associations regarding parental AIDs and offspring T1D matched the genetic correlations between AIDs and T1D. However, with respect to, for example, CD, the genetic correlation analysis based on European GWAS summary statistics, showed a significant shared non-HLA association, but this could not be shown in the pTDT analysis. We also note that the lack of contribution of the non-HLA PGS to T1D risk does not mean that individual genes have no contribution. It is possible that the overall scores diluted the effects of individual regions and that our analysis may yield different results if only the overlap genes were considered. Significant under-transmission of HLA PGS was observed for IBD and MS, consistent with the opposite pleiotropic HLA effects between T1D and MS or IBD in both the HLA haplotype analysis and multi-allelic HLA PGS analysis, while their non-HLA risk variants were not associated with T1D. The overall lack of familial aggregation suggested that this HLA protective effect could be counterbalanced by other non-captured genetic effects or by environmental effects shared within the family.

No diseases exhibited significant over-transmission exclusively for non-HLA PGS. Even among the disease group presenting over-transmission for both HLA and non-HLA, supporting the role of HLA as the primary driver of genetic risk for many AIDs.^42,43^ The interplay of HLA and non-HLA variants on disease susceptibility was complex. A cross-disease genomic analysis of nine AIDs, such as T1D, RA, SLE, suggested that the top prioritized genes of each analyzed AID converged on the same common pathways relevant to T cell activation and signaling, although distinct genes were prioritized across diseases,^44^ which to some extent explained why they were tightly associated with T1D in our analyses.

Most AIDs are more prevalent in women than in men, and disease progression and severity could also differ between the two sexes.^35,45^ T1D is an exception in that a slight majority of the cases (no matter early-onset or adult-onset) are men, and twice as many fathers than mothers of the patients with T1D also have T1D.^35,36,46^ Similar to previous reports, we also observed that among individuals with T1D, men had a higher risk of having offspring with T1D than women, while significant sex-specific associations affecting transmission were overall not observed among parental AIDs. Further, when at least one parent had T1D, especially if it was the father, the male offspring would have a higher risk of developing T1D than the female offspring. To date, the precise cause of a higher rate of T1D transmission from fathers than from mothers has not yet been identified, but potential hypotheses largely centered around in-utero exposure to different aspects of maternal T1D.

Previous studies also showed that people with AIDs, are less likely to have children.^47^ This is likely to reflect multiple factors, including a higher rate of miscarriage amongst women with AIDs, including T1D, amongst whom the rate of miscarriage is 15-30% higher than in the background population.^48–50^ In some cases, individuals with AIDs may choose not to have children due to a fear of transmitting disease to offspring. Being able to estimate early T1D risk trajectories for a couple’s offspring, utilizing their PGS and AID disease status, will equip them with more accurate information to make such decisions around family planning and provide reassurance to those whose offspring have a low predicted risk of T1D. For individuals with offspring predicted to be at high risk of disease, recent advances in T1D research may mean that targeted screening and/or the option of early preventative therapies may be a realistic option.

So far, we are unaware of any previous study using parental PGS to estimate an AID. We introduced a new approach to calculating PGSs for diseases that have a strong HLA signal, by training different models for HLA and non-HLA regions and combining them based on their expected contribution to the disease. We showed that parental PGS for T1D is strongly predictive of T1D risk before the age of 20 and that parental PGS for other AIDs does not help better predict T1D, given the high prediction accuracy already achieved with T1D PRS. We showed that a T1D PGS that represents the genetic risk factors from parents is positively associated with developing T1D in offspring, especially sons. The cumulative risk trajectories start to plateau around the age of 14-16, especially for children with high parental PGS (top 10%), which is consistent with the incidence peak at 10-14 years reported in previous studies.^4–6^ The effect of high parental PGS on T1D risk in children was, in many instances, higher than a diagnosis of T1D in their mother or father. For example, having a mother with T1D resulted in a cumulative T1D risk of 1.49% by age 20, while the top 10% of parents with the highest PGS had a cumulative risk of 2.88%. Considering that only 0.15% of mothers with children had T1D in Finland, the PGS provides additional value on top of family history. We note that using both parental information and expected birth year for T1D risk estimation will generate different results, considering that temporal factors such as shifts in lifestyle and changes in healthcare policies can influence T1D outcomes. Due to the low number of parental T1D cases in the FinnGen cohort (98 out of 8,872), we did not include both parental PGS and family T1D history in one model for estimation. However, our results indicate the intriguing possibility of considering parental PGS in conjunction with clinical diagnoses to inform parents about T1D risk in their offspring and highlight the effectiveness of Full-PGS especially when a couple do not have T1D records.

This study also has several limitations. One limitation was that AID diagnoses for some patients might not be well captured in the healthcare register. For example, the specific ICD-9 code for T1D was not implemented until 1987, although the Care Register for Finnish Health Care began as early as 1969. Similarly, although the 45 years of follow-up were already quite long for parents, it might still be relatively incomplete for some late-onset diseases (e.g., RA). Secondly, rather than being nationally representative, the amalgamated Finnish biobanks represent a collection of biobanks with diverse methods of data collection. Some of the diseases might have a higher prevalence in the Finnish biobanks compared to the nationally representative FinRegistry. And most of our analyses were conducted using the data from Finland. Although these aspects might limit the generalizability of the results, we noted that our findings in the Finnish biobanks were consistent with those we observed from FinRegistry and matched well with previous studies. Thirdly, due to the limited number of parental AID cases and lack of related information in FinnGen, we were not able to consider potential non-genetic effects (e.g., dietary, lifestyle) and random effects for T1D estimation in offspring. Another limitation was that rather than extending the analysis to siblings or other relatives, in this study, we focused on parents and offspring. We note the HLA and non-HLA association patterns with T1D across AIDs would be similar to what we observed here, although the effect sizes would differ. Finally, while environmental factors (e.g., socioeconomic factors, genetic nurture) will impact the development of T1D and, in a broader scenario, seems likely to steer the development of different AIDs among individuals within the same family, our study mainly focused on family history of diseases and genetic factors by controlling these factors with delicate analytical designs.

In conclusion, our results, while confirming the existence of general familial aggregation of AIDs and T1D, highlight a substantial heterogeneity in the impact of different AIDs on T1D. This heterogeneity is partially explained by different genetic effects within and outside the HLA regions, indicating two different transmission patterns regarding shared genetic liabilities in HLA and non-HLA. Overall, genetic effects inside and outside HLA regions are consistent with observational analyses, but in the case of IBD and MS, we revealed an unexpected divergence between the genetic effects and epidemiological observations. Investigating the mechanisms behind these findings may provide valuable insights into the origins of T1D and the etiology of the familial aggregations of AIDs. Moreover, the relative contribution of HLA and non-HLA genetic risk can be leveraged to create PGS that complements a family history of AIDs as a tool to inform a couple about expected T1D risk in their offspring. Our study design can be extended to other projects studying diseases with familial aggregation or caused by pleiotropy.

## Data Availability

Data description for FinRegistry is publicly available on its website (www.finregistry.fi/finnish-registry-data). Access to FinRegistry data at an individual level is possible by submitting a data permit application (https://asiointi.findata.fi). The application includes information on the purpose of data use; the requested data, including the variables, definitions for the target and control groups, and external datasets to be combined with FinRegistry data; the dates of the data needed; and a data utilization plan. The requests will be evaluated on a case-by-case basis. Once approved, the data will be sent to a secure environment Kapseli and can be accessed within the European Economic Area (EEA) and within countries with an adequacy decision from the European Commission. The Finnish biobank data can be accessed through the Fingenious services (https://site.fingenious.fi/en/) managed by FINBB.

## Acknowledgments

FW was funded by the University of Helsinki and the University of Edinburgh joint PhD program in Human Genomics. AG was supported by the European Research Council (ERC) under the European Union’s Horizon 2020 research and innovation programme [grant number 945733], starting grant AI-Prevent. AG was supported by the Academy of Finland (grant no. 323116). This project also received funding from the European Union’s Horizon 2020 Research and Innovation Programme under grant agreement no. 101016775. TT was supported by grants from Folkhälsan Research Foundation, The Academy of Finland (336822, 312072 and 336826), and the University of Helsinki.

We thank the entire FinRegistry and FinnGen team for making the data available for this study. We also acknowledge, Samuel Jones (Institute for Molecular Medicine Finland (FIMM), Finland) for discussion on HLA PGS analysis, Bradley Jermy (FIMM, Finland) for discussion on genetic correlation analysis, Mary P Reeve (FIMM, Finland) for discussion on disease definitions, Om Dwivedi (FIMM, Finland) for discussion on T1D PGS, and Alessio Gerussi (University of Milano-Bicocca, Italy) for discussion on results from a clinical standpoint. The We want to acknowledge the participants and investigators of FinnGen study. The FinnGen project is funded by two grants from Business Finland (HUS 4685/31/2016 and UH 4386/31/2016) and the following industry partners: AbbVie Inc., AstraZeneca UK Ltd, Biogen MA Inc., Bristol Myers Squibb (and Celgene Corporation & Celgene International II Sàrl), Genentech Inc., Merck Sharp & Dohme LCC, Pfizer Inc., GlaxoSmithKline Intellectual Property Development Ltd., Sanofi US Services Inc., Maze Therapeutics Inc., Janssen Biotech Inc, Novartis Pharma AG, and Boehringer Ingelheim International GmbH. Following biobanks are acknowledged for delivering biobank samples to FinnGen: Auria Biobank (www.auria.fi/biopankki), THL Biobank (www.thl.fi/biobank), Helsinki Biobank (www.helsinginbiopankki.fi), Biobank Borealis of Northern Finland (https://www.ppshp.fi/Tutkimus-ja-opetus/Biopankki/Pages/Biobank-Borealis-briefly-in-English.aspx), Finnish Clinical Biobank Tampere (www.tays.fi/en-US/Research_and_development/Finnish_Clinical_Biobank_Tampere), Biobank of Eastern Finland (www.ita-suomenbiopankki.fi/en), Central Finland Biobank (www.ksshp.fi/fi-FI/Potilaalle/Biopankki), Finnish Red Cross Blood Service Biobank (www.veripalvelu.fi/verenluovutus/biopankkitoiminta), Terveystalo Biobank (www.terveystalo.com/fi/Yritystietoa/Terveystalo-Biopankki/Biopankki/) and Arctic Biobank (https://www.oulu.fi/en/university/faculties-and-units/faculty-medicine/northern-finland-birth-cohorts-and-arctic-biobank). All Finnish Biobanks are members of BBMRI.fi infrastructure (www.bbmri.fi). Finnish Biobank Cooperative -FINBB (https://finbb.fi/) is the coordinator of BBMRI-ERIC operations in Finland. The Finnish biobank data can be accessed through the Fingenious® services (https://site.fingenious.fi/en/) managed by FINBB.

## Author contributions

FW, AL, TT, and AG designed the study and wrote the manuscript, with comments from all authors. FW processed the registry data, conducted the statistical analyses, and generated all figures and tables; AL and FW preprocessed the FinRegistry pedigree data, and the FinRegistry team preprocessed all other registry data of FinRegistry; the FinnGen data team preprocessed the FinnGen data; JR processed HLA amino acid data. TT, PV, and the FinnGen clinical team defined disease endpoints; TT, and LA oversaw the project and interpreted the findings from a clinical standpoint. AL, ZY, SJ, and AG advised the registry-based epidemiological analyses; AL, ZY, AG, and RO advised genetic analyses; AL, SK, JR, JP, and TT advised HLA haplotype analyses; AL, ZY and JR advised amino acid analyses. AG, TT, and AL co-supervised the study. MP is the principal investigator of the FinRegistry project. All authors discussed the results, revised the manuscript, and had final responsibility for the decision to submit for publication.

## Ethics statement

FinRegistry is a collaboration project of the Finnish Institute for Health and Welfare (THL) and the Data Science Genetic Epidemiology research group at the Institute for Molecular Medicine Finland (FIMM), University of Helsinki. The FinRegistry project has received approvals for data access from THL (THL/1776/6.02.00/2019 and subsequent amendments), Digital and Population Data Services Agency (VRK/5722/2019–2), Finnish Center for Pension (ETK/SUTI 22003) and Statistics Finland (TK-53–1451-19). The FinRegistry project has received IRB approval from THL (Kokous 7/2019).

The use of the biobank donor samples is in accordance with the biobank consent and meets the requirements of the Finnish Biobank Act 688/2012. Alternatively, separate research cohorts, collected prior the Finnish Biobank Act came into effect (in September 2013) and start of FinnGen (August 2017), were collected based on study-specific consents and later transferred to the Finnish biobanks after approval by Fimea (Finnish Medicines Agency), the National Supervisory Authority for Welfare and Health. Recruitment protocols followed the biobank protocols approved by Fimea. The Coordinating Ethics Committee of the Hospital District of Helsinki and Uusimaa (HUS) statement number for the FinnGen study is Nr HUS/990/2017.

The FinnGen study is approved by Finnish Institute for Health and Welfare (permit numbers: THL/2031/6.02.00/2017, THL/1101/5.05.00/2017, THL/341/6.02.00/2018, THL/2222/6.02.00/2018, THL/283/6.02.00/2019, THL/1721/5.05.00/2019 and THL/1524/5.05.00/2020), Digital and population data service agency (permit numbers: VRK43431/2017-3, VRK/6909/2018-3, VRK/4415/2019-3), the Social Insurance Institution (permit numbers: KELA 58/522/2017, KELA 131/522/2018, KELA 70/522/2019, KELA 98/522/2019, KELA 134/522/2019, KELA 138/522/2019, KELA 2/522/2020, KELA 16/522/2020), Findata permit numbers THL/2364/14.02/2020, THL/4055/14.06.00/2020, THL/3433/14.06.00/2020, THL/4432/14.06/2020, THL/5189/14.06/2020, THL/5894/14.06.00/2020, THL/6619/14.06.00/2020, THL/209/14.06.00/2021, THL/688/14.06.00/2021, THL/1284/14.06.00/2021, THL/1965/14.06.00/2021, THL/5546/14.02.00/2020, THL/2658/14.06.00/2021, THL/4235/14.06.00/2021, Statistics Finland (permit numbers: TK-53-1041-17 and TK/143/07.03.00/2020 (earlier TK-53-90-20) TK/1735/07.03.00/2021, TK/3112/07.03.00/2021) and Finnish Registry for Kidney Diseases permission/extract from the meeting minutes on 4th July 2019.

The Biobank Access Decisions for FinnGen samples and data utilized in FinnGen Data Freeze 11 include: THL Biobank BB2017_55, BB2017_111, BB2018_19, BB_2018_34, BB_2018_67, BB2018_71, BB2019_7, BB2019_8, BB2019_26, BB2020_1, BB2021_65, Finnish Red Cross Blood Service Biobank 7.12.2017 (000-2018), Helsinki Biobank HUS/359/2017, HUS/248/2020, HUS/430/2021 §28, §29, HUS/150/2022 §12, §13, §14, §15, §16, §17, §18, §23, §58 and §59, Auria Biobank AB17-5154 and amendment #1 (August 17 2020) and amendments BB_2021-0140, BB_2021-0156 (August 26 2021, Feb 2 2022), BB_2021-0169, BB_2021-0179, BB_2021-0161, AB20-5926 and amendment #1 (April 23 2020) and it’s modification (Sep 22 2021), BB_2022-0262, BB_2022-0256, Biobank Borealis of Northern Finland_2017_1013, 2021_5010, 2021_5018, 2021_5015, 2021_5015 Amendment, 2021_5023, 2021_5023 Amendment, 2021_5017, 2022_6001, 2022_6006 Amendment, BB22-0067, 2022_0262, Biobank of Eastern Finland 1186/2018 and amendment 22§/2020, 53§/2021, 13§/2022, 14§/2022, 15§/2022, 27§/2022, 28§/2022, 29§/2022, 33§/2022, 35§/2022, 36§/2022, 37§/2022, 39§/2022, 7§/2023, Finnish Clinical Biobank Tampere MH0004 and amendments (21.02.2020 & 06.10.2020), 8§/2021, 9§/2021, §9/2022, §10/2022, §12/2022, 13§/2022, §20/2022, §21/2022, §22/2022, §23/2022, 28§/2022, 29§/2022, 30§/2022, 31§/2022, 32§/2022, 38§/2022, 40§/2022, 42§/2022, 1§/2023, Central Finland Biobank 1-2017, BB_2021-0161, BB_2021-0169, BB_2021-0179, BB_2021-0170, BB_2022-0256, and Terveystalo Biobank STB 2018001 and amendment 25th Aug 2020, Finnish Hematological Registry and Clinical Biobank decision 18th June 2021, Arctic biobank P0844: ARC_2021_1001.

## STAR Methods

### KEY RESOURCES TABLE

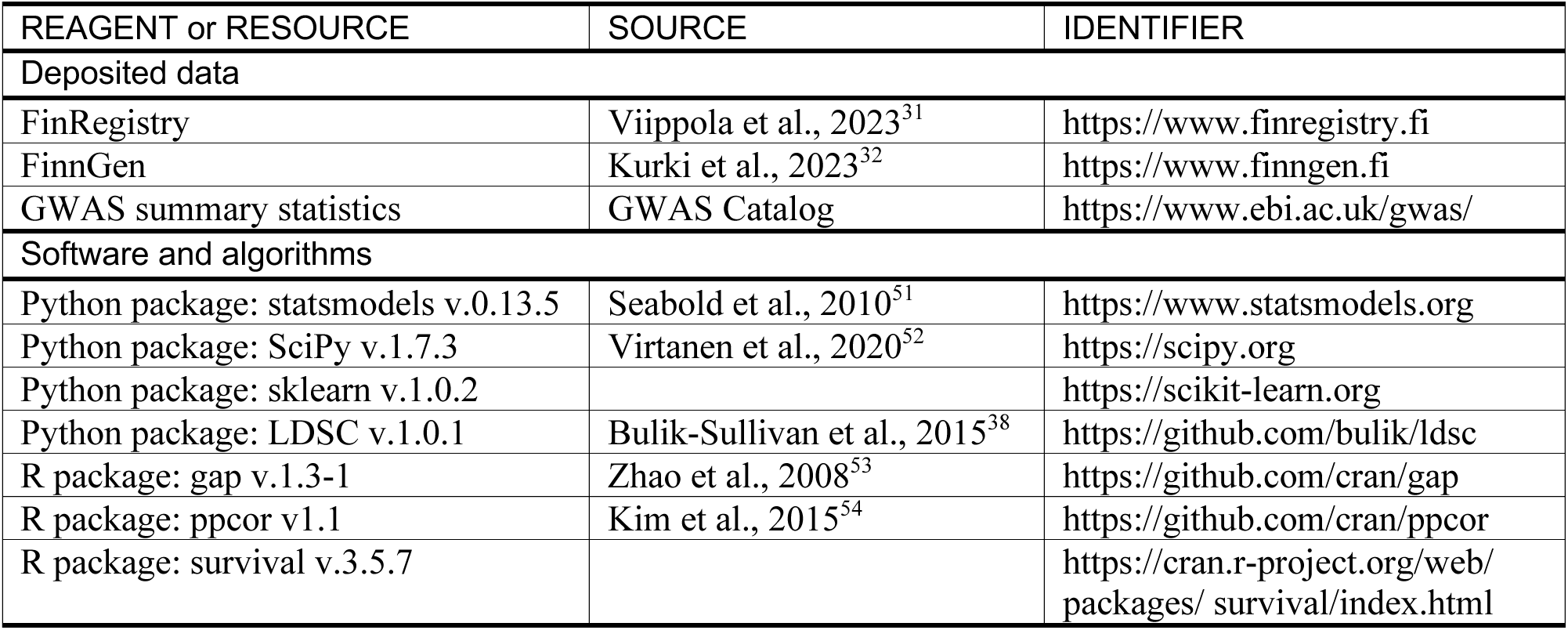

### RESOURCE AVAILABILITY

#### Lead contact

Further information and requests for resources should be directed to and will be fulfilled by the lead contact, Andrea Ganna.

#### Materials availability

This study did not generate new unique reagents.

#### Data and code availability

This work used genotype and phenotype data from FinRegistry (https://www.finregistry.fi) and FinnGen (https://www.finngen.fi). Data description for FinRegistry is publicly available on its website (www.finregistry.fi/finnish-registry-data). Access to FinRegistry data at an individual level is possible by submitting a data permit application (https://asiointi.findata.fi). The application includes information on the purpose of data use; the requested data, including the variables, definitions for the target and control groups, and external datasets to be combined with FinRegistry data; the dates of the data needed; and a data utilization plan. The requests will be evaluated on a case-by-case basis. Once approved, the data will be sent to a secure environment Kapseli and can be accessed within the European Economic Area (EEA) and within countries with an adequacy decision from the European Commission. The Finnish biobank data can be accessed through the Fingenious® services (https://site.fingenious.fi/en/) managed by FINBB. For non-HLA PGS construction, we used GWAS summary statistics from GWAS Catalog (https://www.ebi.ac.uk/gwas/).

Code for the complete analyses is available at https://github.com/dsgelab/parentalAIDs_T1D.

Any additional information required to reanalyze the data reported in this paper is available from the lead contact upon request.

### METHOD DETAILS

#### Data sources

FinRegistry is a national registry database in Finland combining disease registers with comprehensive records on individuals’ demographics, socioeconomic status, death, drug purchases, prescriptions, and administrative information. It includes all residents in Finland who were alive on 1^st^ January 2010, and their first-degree relatives (in total approximately 7.2 million individuals). 572,640 individuals were excluded as they migrated in or out of Finland by 2019, remaining 3,412,326 individuals with both father and mother information.

FinnGen (R11) is a national research project using samples and data from Finnish biobanks, comprising genome-wide genotype data and healthcare registry data for approximately 0.47 million Finnish biobank donors. After excluding 9,196 individuals who had migrated in or out of Finland by 2019, removing 10,588 individuals without imputed HLA data and 8,627 individuals who have other missing values in the cohort, we included 3,668 T1D cases and 441,602 controls and observed lifetime prevalence and number of cases in FinnGen for each selected AID. The study population features slightly more women (N=251,294, 56.4%).

By using the datasets from the same country, we alleviate concerns about potential bias caused by differences in genetic background, healthcare system, or socio-cultural context.

#### Finnish population-based case-control study

The epidemiology association between AIDs in parents and T1D in their offspring was investigated by a case-control study within FinRegistry. To include most of the individuals and their parents in this analysis, we selected all individuals born in Finland between 1960 and 1999 who had both father’s and mother’s information available. The follow-up period started either from the date they were born or from the year the codes for our selected diseases were included in national patient register and ended on 31 December 2019, which is the latest follow-up date in FinRegistry. We then restricted our analysis to offspring whose father was born between 1917 and 1976 and whose mother was born between 1922 and 1976. Finally, we removed those individuals who died during the follow-up period.

T1D cases were individuals diagnosed with T1D before the age of 40. For each trio with T1D-affected children, three controls without T1D diagnosis were matched by following variables: sex, birth year (five-year bin, 8 levels), and birthplace for the child (19 levels), birth year of father (five-year bin, 12 levels), birth year of mother (five-year bin, 11 levels), and family size (represented by the number of siblings: 0, 1, 2, 3, >=4). The differences in socio-demographic characteristics between cases and controls, including socio-economic status, marital status, number of offspring, occupation, education, and mother tongue, were further examined to ensure that the two groups were comparable. Cohen’s D^55^ (≥0.2) for ordinal characteristics such as number of siblings and number of children and chi-squared test (P<0.05) for categorical variables such as education and marital status were used to test whether the differences are minor.

Exposures of interest were all the parental endpoints related to AIDs mainly defined by Finnish versions 8-10 of ICD. AIDs with fewer than 50 cases among parents in the final study population in the case-control study were filtered out from the list. We then excluded AID codes that were subtypes of other codes (Supplementary notes 2.1).

A conditional logistic regression, adjusted by individual’s birth year, birth year of father, birth year of mother, and the exact number of siblings, was applied to estimate the effect of having a given AID in parents on the risk of developing T1D in offspring. Bonferroni correction was applied, and only the diseases reached significant associations (P<=0.05/26) were included for further analyses. To examine whether the identified associations are shared between father and mother, we used conditional logistic regression stratified by maternal AIDs status and paternal AIDs status. We also investigated whether parental AID status can influence the onset age of disease in offspring by regressing the age at onset of T1D in offspring on the AID status in their parents.

#### Shared genetic components at a population level

To explore, to what extent, the familial aggregations between AIDs and T1D identified in the population-based case-control study are contributed by genetic factors, we conducted four analyses in both HLA regions and non-HLA regions. Considering the strong associations between HLA and AIDs,^3^ we first performed an HLA haplotype analysis and an HLA-based PGS analysis using imputed HLA alleles. We also converted the HLA alleles to HLA amino acids and conducted AA analysis. Then, we performed a genetic correlation analysis using variants in non-HLA regions.

##### HLA haplotype and amino acid analyses

The HLA alleles were imputed with a population-specific reference panel.^56^ We considered all the 187 unique alleles on 10 classical HLA genes in class I and II regions for analysis, including 27 alleles for *HLA-A*, 40 for *HLA-B*, 23 for *HLA-C*, 24 for *HLA-DPB1*, 15 for *HLA-DQA1*, 16 for *HLA-DQB1*, 33 for *HLA-DRB1*, and 3 each for *HLA-DRB3*, *HLA-DRB4*, and *DRB5*. According to previous studies,^7,8,34^ several HLA haplotypes in Class II have been significantly associated with an increased risk of developing T1D and other AIDs. To understand whether HLA genetic liability to T1D is associated with other AIDs, we conducted a haplotype analysis (Figure S2.4.1) and only considered possible haplotypes from gene combination *HLA-DRB1-DQA1-DQB1*. We first removed individuals with any ambiguous genotypes for the given haplotype. We define an ambiguous genotype as a genotype with imputed allele dosage in [0.4, 0.6] or [1.4,1.6]. Next, we extracted the potential haplotypes using the expectation-maximization algorithm and removed those haplotypes whose frequency is less than 0.005. We identified the most prevalent HLA haplotypes that were either positively or negatively associated with T1D using a haplotype score test (P<1.0 ×10^-10^), a statistical test to calculate the contribution of a specific haplotype on a certain phenotype when the linkage phase is unknown.^57^ Bonferroni corrections were applied for haplotype selection. Afterward, a multivariable logistic regression was built to estimate the associations between the remaining haplotypes and AID status, adjusting for age, sex, and the first ten principal components (PCs).

To better understand the HLA pleiotropy across the AIDs, we further mapped HLA alleles, including those of genes DRB1, DQA1, and DQB1, to protein sequences and tested their associations with the AIDs. We used the data from IPD-IMGT/HLA Database (version: 3.58.0) for the mapping and then assured every individual has numerical dosage information at each amino acid position in the protein alignment.^58^ We then ran the logistic regression model with the same settings as the HLA allele-based analyses.

##### HLA multi-allelic score

To tackle the complex LD structure and high polymorphism of HLA alleles, we developed a novel method to construct PGS specific to HLA genes for T1D and other AIDs. We considered all individuals of genotyped family trios as our target population and the rest as a training set. To generate proper weights for analyzed HLA alleles, we applied weighted ridge classifiers with optimal regularization strength, which imposed a penalty on the size of the coefficients. After PGS normalization, we first measured the partial correlation between each AID and its PGS. We dropped all the AIDs whose partial correlation with PGS for the AIDs themselves was lower than 2%, to filter out diseases with low sample sizes or with weak HLA signals. We then conducted logistic regression to examine the association between PGS and T1D. We also compared the weighted ridge classifiers to other ensemble methods, including lasso regression and elastic net, and found that the former showed higher increment of pseudo-R^2^ and AUC for most of the analyzed AIDs (Table S3.3.3.4; Table S3.3.3.5). Bonferroni correction was applied. For covariates, we adjusted for age, sex, and the first ten PCs.

##### Genome-wide genetic correlation with HLA regions excluded

To evaluate the genetic overlap between T1D and other AIDs due to shared non-HLA effects, we estimated genetic correlation using LDSC, considering common variants across the whole genome except for the extended HLA regions (chromosome 6: 25–34 Mb).^38,39^ For summary statistics, we used GWAS of European ancestry from the studies with largest sample size (Table S2.3.1). For sensitivity analyses, we further examined whether the estimated genetic correlations changed significantly when excluding all SNPs on chromosome 6. We considered HapMap3^59^ SNPs and constructed LD structures with European ancestry samples from the 1000 Genomes project.^60^

#### Intergeneration cross-trait inheritance with polygenic transmission disequilibrium test

To understand the transmission of parental AIDs to T1D in offspring, we further estimated the contribution of parental transmitted and non-transmitted genetic liability to T1D in offspring for both HLA and non-HLA genes using data from FinnGen. While offspring are expected to receive, on average, half of their genetic materials from each parent, some will over or under-inherit alleles associated with the disease from parents, which will have an impact on their disease risk. We applied a polygenic transmission disequilibrium test (pTDT) to assess whether the mean of the offspring PGS distribution is consistent with its parentally derived expected value.^40^

For non-HLA PGS, we applied PRS-CS and used the same GWAS summary statistics as genetic correlation analysis (Table S2.3.2). We included 12,563 genotyped trios for which both parents and at least one offspring were directly genotyped. We considered HLA PGSs and non-HLA PGSs that were robustly associated with the corresponding AID (P<0.05/19 and |ρ|>=2%). This resulted in ten diseases with both reliable HLA PGS and non-HLA PGS: T1D, HYPO, CD, RA, SLE, sarcoidosis, psoriasis, IBD, MS, and AS, as well as an additional six diseases for HLA PGS (B12A, HYPER, PBC, mixed connective tissue disease, Sjögren’s syndrome, and ADDISON) (Table 3.3.3.2; Table S3.4.1).

We removed genotyped families with affected parents and then subtracted mid-parent PGS for that AID from offspring PGS for the same disease. A significant over-transmission of PGS for an AID from parents to their offspring, suggesting a positive correlation between this AID and T1D. Conversely, significant under-transmission indicates genetic opposite effects between T1D and this AID. If the transmission pattern is neutral, it reflects a lack of correlation.

### Impact of parental AIDs on cumulative risk of T1D in children

To assess how well parental PGSs could be linked to T1D in offspring, we designed several analyses to test the associations and predicting abilities. Considering the prediction performance of PGS for parental AIDs, we only focused on those AIDs that showed significance from previous PGS analyses in either HLA or non-HLA regions. We introduced a new approach, Full-PGS, to construct PGS combining separately generated HLA and non-HLA PGSs (Supplementary notes 2.5).

For each parental AID, we constructed a mid-parent Full-PGS using HLA PGS and non-HLA PGS considering their ratio for cross-disease prediction and examined the association with T1D in offspring. For comparisons, we ran additional models including only parental diagnosis, only mid-parent PGS calculated by a traditional PGS approach that takes genome-wide variants altogether for calculation, e.g., PRS-CS,^41^ or both HLA PGS and non-HLA PGS from our previous analyses, assumed to be at least as good as Full-PGS. We used AUC as a primary metric to evaluate model performances. To understand to what extent the additive value of parental PGS for other AIDs can be added to a T1D prediction model, we then compare a model with all mid-parent PGSs to a model with only mid-parent T1D PGS. We evaluated the two models by comparing the AUCs among 12,563 genotyped trios in FinnGen. To avoid overfitting, the analyses were done with five-fold cross-validation.

We then conducted a Cox proportional hazards model to assess the T1D cumulative incidence risk in offspring before age 20, adjusting for birth years and the first 10 PCs of offspring. To maximize the statistical power, we considered all genotyped trios with children born between 1960 and 2010. Among the 8,827 trios we analyzed, 1,035 had at least one offspring with T1D (11.7%). We then grouped the trios by dividing the parental Full-PGS percentile into 0-50^th^, 50-90^th^, 90-99^th^, and 99-100^th^ and estimated cumulative risk within each group given the mean of children’s birth year and PCs. We also stratified the model by the sex of offspring. To make the result generalizable, we calibrated the FinnGen results based on the FinRegistry’s nationwide T1D prevalence among all children born between 1960-2010 (mean_birth year_=1984; N=3,048,812; prevalence=0.6%).

In FinRegistry, we applied a similar design and considered four groups: 1) parents without T1D, 2) maternal T1D only, 3) paternal T1D only, and 4) both parents with T1D among the whole study population. For the analyses excluding children with parental T1D, we considered four groups: 1) parents without AID, 2) only the mother had AID(s), 3) only the father had AID(s), and 4) both parents had AID(s). The parental AIDs in this analysis covered all the significant associations identified from the previous analyses.

