## Supplementary notes for "Effects of parental autoimmune diseases on type 1 diabetes in offspring can be partially explained by HLA and non-HLA polymorphisms"

#### Table of Contents


**1. Supplementary background**

**2. Supplementary methods**

- 2.1. Disease definition and source codes
- 2.2. Inclusion and exclusion criteria of the matched case-control study
- 2.3. GWAS summary statistics for LDSC and PGS analyses
- 2.4. Study design for HLA analysis
  - 2.4.1. Design of the HLA haplotype analysis
  - 2.4.2. Design of the HLA polygenic risk analysis
- 2.5. Full-PGS construction

**3. Supplementary results**

- 3.1. Basic statistics and demographic data
  - 3.1.1. Basic estimates in FinRegistry and FinnGen
  - 3.1.2. Socio-demographic characteristics in FinRegistry
- 3.2. Registry-based analyses
  - 3.2.1. Matched case-control study
  - 3.2.2. Association between parental AIDs and T1D in offspring by sex
  - 3.2.3. Association between parental AIDs and T1D onset age in offspring by sex
  - 3.2.4. Sensitivity analyses
- 3.3. Shared genetic background in population
  - 3.3.1. Analyses in HLA regions – haplotype analysis
  - 3.3.2. Analyses in HLA regions – amino acid analysis
  - 3.3.3. Analyses in HLA regions – PGS Analysis
  - 3.3.4. Analyses in non-HLA regions – LDSC
- 3.4. Inter-generation cross-trait transmission
  - 3.4.1. Analyses in non-HLA regions – PGS analysis
  - 3.4.2. Polygenic transmission disequilibrium tests
- 3.5. Contribution ratio between HLA PGS and non-HLA PGS
  - 3.5.1. Associations between PGSs for AIDs and T1D in population
  - 3.5.2. Contribution ratio between HLA PGS and non-HLA PGS
- 3.6. Construction of Full-PGS
  - 3.6.1. Results for AID prediction using PGS for AID
  - 3.6.2. Results for T1D prediction using PGS for AID
  - 3.6.3. Results for T1D prediction in offspring using mid-parent PGS for AID
  - 3.6.4. PTDT comparisons between Full-PGS and PRS-CS

**4. Supplementary discussion**

**5. Reference**

### 1. Supplementary Background

**Table S1: Case reports, clinical studies and population-based studies reporting either co-occurrence or familial aggregation of the listed autoimmune diseases among patients with type 1 diabetes**

| Disease | Co-occurrence | Familial aggregation | |
|---|---|---|---|
| | | Clinical study | Genetic study |
| Type 1 diabetes | N/A | Sipetić et al. [8]<br>Hanukoglu et al. [13]<br>Anaya et al. [19]<br>Hemminki et al. [24] **<br>Kishi et al. [26]<br>Olamoyegun et al. [43]<br>Yen et al. [46] **<br>Kang et al. [47] | Douek et al. [10]<br>Lambert et al. [15]<br>Erlich et al. [22]<br>Bronson et al. [25]<br>Parkkola et al. [34] |
| Adrenocortical insufficiency | Barker et al. [17]<br>Van den Driessche et al. [27]<br>Kota et al. [33]<br>Conrad et al. [49] ** | Hemminki et al. [24] ** | Wägner et al. [30]<br>Parkkola et al. [35] |
| Autoimmune hyperthyroidism | Gray et al. [5]<br>De Block et al. [23]<br>Van den Driessche et al. [27]<br>Kota et al. [33]<br>Kahaly et al. [41]<br>Kang et al. [47]<br>Conrad et al. [49] ** | Anaya et al. [19]<br>Hemminki et al. [24] **<br>Ganji et al. [42]<br>Yen et al. [46]<br>Kang et al. [47] | Wägner et al. [30]<br>Parkkola et al. [35] |
| Autoimmune hypothyroidism | Samantaray et al. [3]<br>Gray et al. [5]<br>Kordonouri et al. [9]<br>Hanukoglu et al. [13]<br>De Block et al. [23]<br>Van den Driessche et al. [27]<br>Nagy et al. [28]<br>Kota et al. [33]<br>Karavanaki et al. [37]<br>Kahaly et al. [41]<br>Kang et al. [47]<br>Conrad et al. [49] ** | Bottazzo et al. [4]<br>Hanukoglu et al. [13]<br>Anaya et al. [19]<br>Hemminki et al. [24] **<br>Karavanaki et al. [37]<br>Yen et al. [46] **<br>Kang et al. [47] | Wägner et al. [30]<br>Parkkola et al. [35] |
| Autoimmune hemolytic anemia | Masood et al. [36] | Hemminki et al. [24] | |
| Allergic purpura | D'Souza et al. [38] | | |
| Vitamin B12 deficiency anaemia | Hanukoglu et al. [13]<br>De Block et al. [23]<br>Van den Driessche et al. [27] | Bottazzo et al. [4]<br>Sipetić et al. [8]<br>Anaya et al. [19]<br>Hemminki et al. [24] ** | Wägner et al. [30]<br>Parkkola et al. [35] |
| Idiopathic thrombocytopenic purpura | De Block et al. [23] | Hanukoglu et al. [13]<br>Hemminki et al. [24] | |
| Sarcoidosis | Benmelouka et al. [45] | Hemminki et al. [24] ** | |
| Primary biliary cholangitis | Gazzaruso et al. [12]<br>Johnson et al. [44] | Anaya et al. [19]<br>Hemminki et al. [24] ** | |
| Coeliac disease | Walker-Smith et al. [1]<br>Thain et al. [2]<br>Hanukoglu et al. [13]<br>Sanchez-Albisua et al. [16]<br>Goh et al. [20]<br>Van den Driessche et al. [27] | Sipetić et al. [8]<br>Hanukoglu et al. [13]<br>Anaya et al. [19]<br>Hemminki et al. [24] **<br>Ganji et al. [42]<br>Kang et al. [47] | Wägner et al. [30]<br>Parkkola et al. [35] |

| | | | |
|---|---|---|---|
| | Zeglaoui et al. [29]<br>Bhadada et al. [31]<br>Franzese et al. [32]<br>Kota et al. [33]<br>Kang et al. [47]<br>Conrad et al. [49] ** | | |
| Inflammatory bowel disease | Kang et al. [47]<br>Conrad et al. [49] * | Sipetić et al. [8]<br>Hemminki et al. [24] **<br>Yen et al. [46] *<br>Kang et al. [47] | Wägner et al. [30] |
| IgA nephropathy | Orfila et al. [7] | | |
| Ankylosing spondylitis | | Hemminki et al. [24] * | |
| Mixed connective tissue disease | | Anaya et al. [19] | Parkkola et al. [35] |
| Rheumatoid arthritis | Hanukoglu et al. [13]<br>Nagy et al. [28]<br>Kang et al. [47]<br>Conrad et al. [49] ** | Hanukoglu et al. [13]<br>Anaya et al. [19]<br>Hemminki et al. [24] **<br>Kang et al. [47]<br>Yen et al. [46] | Wägner et al. [30]<br>Parkkola et al. [35] |
| Sjögren syndrome | Binder et al. [6]<br>Shimomura et al. [14] | Anaya et al. [19]<br>Hemminki et al. [24]<br>Yen et al. [46]<br>Kuo et al. [39] ** | Parkkola et al. [35] |
| Systemic sclerosis | Zeglaoui et al. [29] | Hemminki et al. [24] | Parkkola et al. [35] |
| Wegener granulomatosis | | Hemminki et al. [24] ** | |
| Systemic lupus erythematosus | Zeglaoui et al. [29]<br>Kota et al. [33]<br>Masood et al. [36] | Anaya et al. [19]<br>Hemminki et al. [24] **<br>Yen et al. [46] | Wägner et al. [30]<br>Parkkola et al. [35] |
| Guillain-Barre syndrome | Hanukoglu et al. [13] | Hanukoglu et al. [13] | |
| Multiple sclerosis | Marrosu et al. [48] ** | Nielsen et al. [18]<br>Anaya et al. [19]<br>Hemminki et al. [24]<br>Marrosu et al. [48] ** | Wägner et al. [30]<br>Parkkola et al. [35] |
| Myasthenia gravis | Wakata et al. [21]<br>Kota et al. [33]<br>Fang et al. [40] ** | Hanukoglu et al. [13]<br>Hemminki et al. [24] | Wägner et al. [30] |
| Alopecia areata | | Sipetić et al. [8]<br>Hanukoglu et al. [13] | |
| Psoriasis | Hanukoglu et al. [13]<br>Masood et al. [36]<br>Kang et al. [47] | Sipetić et al. [8]<br>Hanukoglu et al. [13]<br>Anaya et al. [19]<br>Hemminki et al. [24]<br>Yen et al. [46]<br>Kang et al. [47] | Wägner et al. [30] |
| Vitiligo | Hanukoglu et al. [13]<br>Van den Driessche et al. [27]<br>Masood et al. [36]<br>Conrad et al. [49] * | Alkhateeb et al. [11]<br>Hanukoglu et al. [13]<br>Anaya et al. [19] | Wägner et al. [30] |

The listed studies indicate that at least one case of co-occurrence or familial aggregation was mentioned. For those studies using large population-based registry data, *: P-value<0.05, **: P-value<0.01

### 2. Supplementary methods

#### 2.1 Disease definition and source codes

We used the disease endpoints as defined by clinical expert groups of the FinnGen study based on the Finnish version International Classification of Diseases (ICD) codes of version 8 (1969-1986), 9 (1987-1995), and 10 (1996-2019). More detailed disease definitions can be found from https://risteys.finregistry.fi/ searching with the FinRegistry code names. The numbers of parental autoimmune disease (AID) cases (Np) were calculated from study population and diseases were filtered according to these numbers (Np<50).

#### Table S2.1: Definition of disease endpoints through ICD-8, ICD-9, and ICD-10

| | | International Classification of Diseases | | | | |
|---|---|---|---|---|---|---|
| **Disease** | **Abbreviation** | **ICD-10** | **ICD-9** | **ICD-8** | $N_p$ | **Delete reason** |
| Type 1 diabetes | T1D | E10 (age < 40) | 250 (age < 40) | 250 (age < 40) | 385 | |
| Adrenocortical insufficiency | ADDISON | E27[1-4] | 2554 | 2551 | 166 | |
| Autoimmune hyperthyroidism | HYPER | E05[0\|9] | 2420 | 2420 | 635 | |
| Autoimmune hypothyroidism | HYPO | E03[8-9] | 244[8-9] | 244 | 11485 | |
| Autoimmune hemolytic anemia | AIHA | D591 | 2830 | 28390 | 55 | |
| Allergic purpura | | D690 | 2870 | 2870 | 154 | |
| Vitamin B12 deficiency anaemia | B12A | D51 | 281[0-1] | 281[0-1] | 962 | |
| Idiopathic thrombocytopenic purpura | ITP | D693 | 2873A | 28710 | 186 | |
| Sarcoidosis | | D86 | 135 | 135 | 1104 | |
| Primary biliary cholangitis | PBC | K743 | N/A | N/A | 157 | |
| Coeliac disease | CD | K900 | 5790A | N/A | 1129 | |
| Inflammatory bowel disease | IBD | K50\|K51 | 555\|556 | 5630\|5631\|569 | 2221 | |
| IgA nephropathy | | N082&D8980 | N/A | N/A | 89 | |
| Ankylosing spondylitis | AS | M45 | 7200 | 7124 | 680 | |
| Mixed connective tissue disease | MCTD | M359 | 7109 | 73499 | 374 | |
| Rheumatoid arthritis | RA | M0[5-6] | 714[0-2] | 712[1-3] | 3224 | |
| Sjögren syndrome | SjS | M350 | 7102 | 73490 | 585 | |
| Systemic sclerosis | SS | M34 | 7101 | 7340 | 118 | |
| Wegener granulomatosis | WG | M313 | 4464 | 4462 | 86 | |
| Systemic lupus erythematosus | SLE | M32 | 7100 | 7431 | 210 | |
| Guillain-Barre syndrome | GBS | G610 | 3570 | 35401 | 120 | |
| Multiple sclerosis | MS | G35 | 340 | 34099 | 461 | |
| Myasthenia gravis | MG | G700 | 3580 | 7330 | 103 | |
| Alopecia areata | AA | L63 | 7040B\|7040C | 70400 | 175 | |

| | | | | | |
|---|---|---|---|---|---|
| Psoriasis | L40 | 6961, 6960A, 6968X | 6869[3\|6]\|696[00\|10\|19] | 2486 | |
| Vitiligo | L80 | 7090F | 70905 | 65 | |
| Relapsing polychondritis | M94.1 | N/A | N/A | 14 | $N_p$<50 |
| Dermatopolymyositis | M33 | N/A | N/A | 91 | Overlap with CD |
| Microscopic polyangiitis | M31.7 | N/A | N/A | 17 | $N_p$<50 |
| Churg-Strauss | M30.1 | 4460B | N/A | 18 | $N_p$<50 |
| Behçet disease | M35.2 | 1361 | N/A | 15 | $N_p$<50 |
| Hypersensitivity angiitis | M31.0 | 4462A | 4461 | 28 | $N_p$<50 |
| Rheumatic fever incl heart disease | I0[0-2\|5-9] | 39[0-8] | 39[0-8] | 262 | Unclear definition |
| Acute disseminated encephalitis | G04.0# | 3235A | 32300 | 13 | $N_p$<50 |
| Other acute disseminated demyelination | G36 | 3410 | 34101 | 45 | $N_p$<50 |
| Narcolepsy and cataplexy | G47.4 | 347[0-1] | 34700 | 30 | $N_p$<50 |
| Autoimmune polyglandular failure | E31.0[0-1\|8] | 2851 | N/A | 9 | $N_p$<50 |
| Graves ophthalmopathy | H06.2*E05.9 | N/A | N/A | 100 | Subtype of HYPER |
| Autoimmune hepatitis | K75.4 | N/A | N/A | 0 | $N_p$<50 |
| Drug-induced autoimmune haemolytic anaemia | D59.0 | 2830B | 28391 | 8 | $N_p$<50 |
| Disorders of myoneural junction and muscle in diseases classified elsewhere | G73* | N/A | N/A | 28 | $N_p$<50 |
| Other demyelinating diseases of the central nervous system | G37 | 341[1-9] | 3410[2-9] | 197 | Unclear definition |
| Anterior Iridocyclitis | H20.0, H20.1 | 3640, 3641 | 36400, 36402 | 1770 | Unclear definition |
| Ulcerative colitis | K51 | 556 | 5631\|569 | 1747 | Subtype of IBD |
| Crohn disease | K50 | 555 | 5630 | 727 | Subtype of IBD |
| Pemphigoid | L12 | N/A | N/A | 144 | Unclear definition |
| Dermatitis herpetiformis | L13.0 | 6940A\|6942 | 693 | 152 | Overlap with CD |
| Henoch-Schönlein purpura nephritis | N08.2*D69.0 | N/A | N/A | 17 | $N_p$<50 |
| Autoimmune thyroiditis | E06.3 | 2452 | N/A | 136 | Unclear definition |
| Linear scleroderma | L94.1 | N/A | N/A | <5 | $N_p$<50 |

**Figure S2.2: Inclusion and exclusion criteria of the matched case-control study**

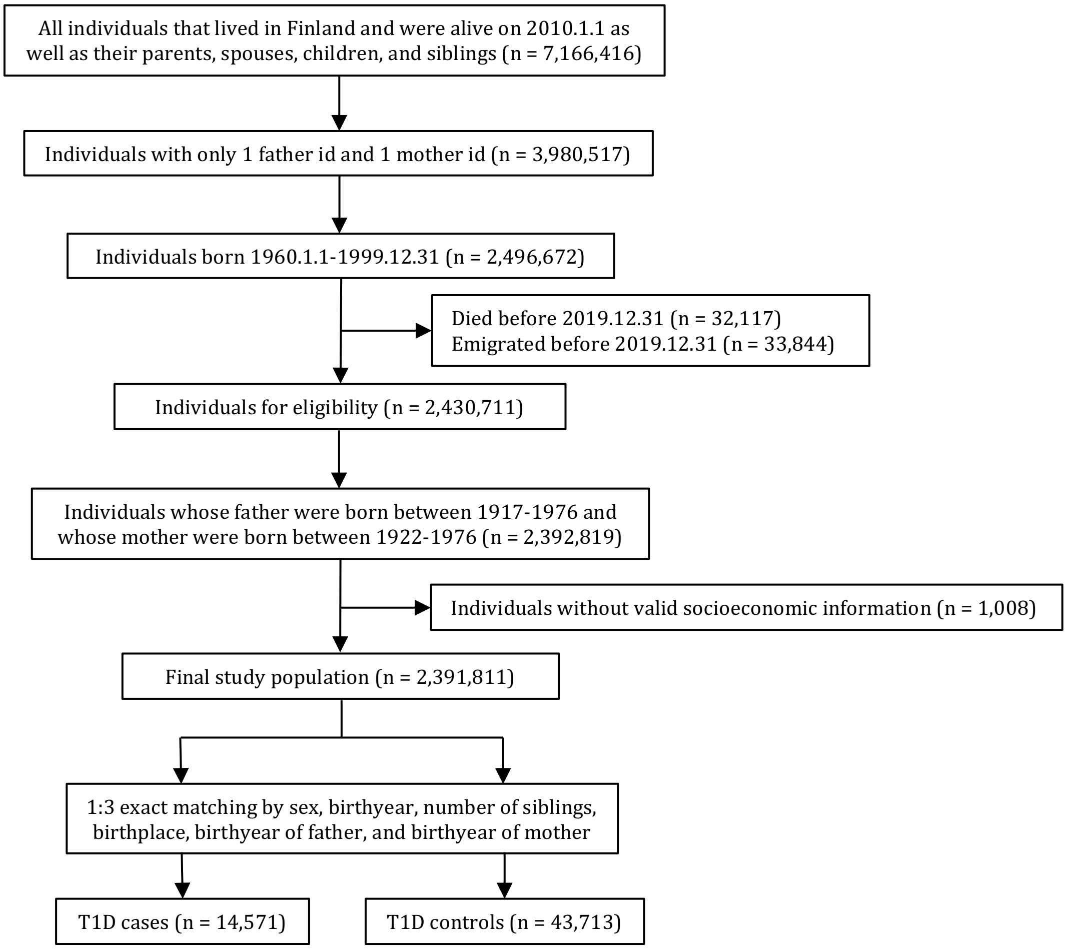

#### 2.3 GWAS summary statistics for LDSC and PGS analyses

For each autoimmune disease, we first searched recent summary statistics with the largest sample size from European ancestry via https://www.ebi.ac.uk/gwas/. If nothing could be found there, we then tried to obtain the statistics from meta-analysis on FinnGen R11, UK Biobank, and Estonia via https://metaresults-est-ukbb.finngen.fi, or meta-analysis on FinnGen R11 and UK Biobank via https://metaresults-ukbb.finngen.fi, or analysis on FinnGen R11 via https://results.finngen.fi.

**Table S2.3.1: The sources of GWAS summary statistics regarding different autoimmune diseases that were used to perform linkage disequilibrium score regression (LDSC) analysis**

To construct non-HLA PGS in genotyped trios or full PGS in imputed trios, for each AD, we extracted publicly accessible GWAS summary statistics with the largest sample size from European ancestry. Cases/controls refer to the number of individuals affected/not affected with the disease in question.

| Disease | Source | Year | Cases | Controls |
|---|---|---|---|---|
| Type 1 diabetes | https://doi.org/10.1038/s41588-021-00880-5 | 2021 | 22 153 | 37 374 |
| Adrenocortical insufficiency | FinnGen R11 | | 1 116 | 434 894 |
| Autoimmune hyperthyroidism | FinnGen R11 + UK Biobank + Estonia | | 4 578 | 1 019 242 |
| Autoimmune hypothyroidism | FinnGen R11 + UK Biobank + Estonia | | 87 043 | 877 215 |
| Autoimmune hemolytic anemia | FinnGen R11 | | 348 | 452 718 |
| Allergic purpura | FinnGen R11 | | 1 031 | 446 531 |
| Vitamin B12 deficiency anaemia | FinnGen R11 + UK Biobank + Estonia | | 9 164 | 989 292 |
| Idiopathic thrombocytopenic purpura | FinnGen R11 | | 968 | 446 531 |
| Sarcoidosis | FinnGen R11 + UK Biobank + Estonia | | 7 031 | 1 062 340 |
| Primary biliary cholangitis | FinnGen R11 + UK Biobank + Estonia | | 1 253 | 836 368 |
| Coeliac disease | https://doi.org/10.1038/ng.543 | 2010 | 4 533 | 10 750 |
| Inflammatory bowel disease | https://doi.org/10.1038/ng.3760 | 2017 | 25 042 | 34 915 |
| IgA nephropathy | FinnGen R11 + UK Biobank | | 944 | 873 331 |
| Ankylosing spondylitis | https://doi.org/10.1038/ng.2667 | 2013 | 9 069 | 13 578 |
| Mixed connective tissue disease | FinnGen R11 | | 2 190 | 451 543 |
| Rheumatoid arthritis | http://dx.doi.org/10.1136/annrheumdis-2020-219065 | 2020 | 22 628 | 288 664 |
| Sjögren syndrome | FinnGen R11 + UK Biobank + Estonia | | 4 323 | 1 047 524 |
| Systemic sclerosis | https://doi.org/10.1038/s41467-019-12760-y | 2019 | 9 095 | 17 584 |
| Wegener granulomatosis | FinnGen R11 | | 486 | 439 424 |
| Systemic lupus erythematosus | https://doi.org/10.1038/ng.3434 | 2015 | 7 219 | 15 991 |
| Guillain-Barre syndrome | FinnGen R11 | | 489 | 445 865 |
| Multiple sclerosis | https://doi.org/10.1038/nature10251 | 2011 | 9 772 | 17 376 |
| Myasthenia gravis | https://doi.org/10.1073/pnas.2108672119 | 2022 | 1 873 | 36 370 |
| Alopecia areata | FinnGen R11 + UK Biobank + Estonia | | 3 384 | 973 626 |
| Psoriasis | FinnGen R11 + UK Biobank + Estonia | | 34 530 | 1 029 934 |
| Vitiligo | https://doi.org/10.1038/ng.3680 | 2016 | 4 680 | 39 586 |

#### Table S2.3.2: The sources of GWAS summary statistics regarding different autoimmune diseases that were used to construct non-HLA PGS and full PGS.

To construct non-HLA PGS in genotyped trios or full PGS in imputed trios, we used GWAS summary statistics excluding FinnGen. After removing seven AIDs using only statistics from FinnGen, we used the same source of GWAS summary statistics by removing that from FinnGen in **Table S2.3.1** for the rest 19 AIDs. Non-HLA PGSs were constructed using the GWAS summary statistics excluding HLA regions. For coeliac disease, the variants in HLA regions were not available in the GWAS summary statistics (**Table S2.3.1**), we used meta-analysis of UK biobank and Estonia biobank instead for full PGS construction. In the last column of **Table S2.3.2**, each row specifies which analysis uses the corresponding data source. "Non-HLA" indicates non-HLA construction. "Full" is full PGS construction.

| Disease | Source | Year | Cases | Controls | Analysis |
|---|---|---|---|---|---|
| Type 1 diabetes | https://doi.org/10.1038/s41588-021-00880-5 | 2021 | 22153 | 37 374 | non-HLA; full |
| Autoimmune hyperthyroidism | UK Biobank + Estonia | | 1141 | 568 946 | non-HLA; full |
| Autoimmune hypothyroidism | UK Biobank + Estonia | | 37988 | 547 440 | non-HLA; full |
| Vitamin B12 deficiency anaemia | UK Biobank + Estonia | | 5182 | 556 285 | non-HLA; full |
| Sarcoidosis | UK Biobank + Estonia | | 2177 | 615 817 | non-HLA; full |
| Primary biliary cholangitis | UK Biobank + Estonia | | 562 | 498 082 | non-HLA; full |
| Coeliac disease | https://doi.org/10.1038/ng.543 | 2010 | 4533 | 10 750 | non-HLA |
| Coeliac disease | UK Biobank + Estonia | | 4533 | 10 750 | full |
| Inflammatory bowel disease | https://doi.org/10.1038/ng.3760 | 2017 | 25042 | 34 915 | non-HLA; full |
| IgA nephropathy | UK Biobank | | 218 | 420 324 | non-HLA; full |
| Ankylosing spondylitis | https://doi.org/10.1038/ng.2667 | 2013 | 9069 | 13 578 | non-HLA; full |
| Rheumatoid arthritis | http://dx.doi.org/10.1136/annrheumdis-2020-219065 | 2020 | 22628 | 288 664 | non-HLA; full |
| Sjögren syndrome | UK Biobank + Estonia | | 1342 | 608 100 | non-HLA; full |
| Systemic sclerosis | https://doi.org/10.1038/s41467-019-12760-y | 2019 | 9095 | 17 584 | non-HLA; full |
| Systemic lupus erythematosus | https://doi.org/10.1038/ng.3434 | 2015 | 7219 | 15 991 | non-HLA; full |
| Multiple sclerosis | https://doi.org/10.1038/nature10251 | 2011 | 9772 | 17 376 | non-HLA; full |
| Myasthenia gravis | https://doi.org/10.1073/pnas.2108672119 | 2022 | 1873 | 36 370 | non-HLA; full |
| Alopecia areata | UK Biobank + Estonia | | 3384 | 973 626 | non-HLA; full |
| Psoriasis | UK Biobank + Estonia | | 34530 | 1 029 934 | non-HLA; full |
| Vitiligo | https://doi.org/10.1038/ng.3680 | 2016 | 4680 | 39 586 | non-HLA; full |

#### 2.4 Study design for HLA analyses

There are two analyses in this section. We first conducted a haplotype based GWAS in HLA region to understand how T1D correlated with other AIDs via HLA haplotypes. Then we investigated whether HLA genetic liability to other ADs have a causal effect on T1D using our novel HLA PGS method.

#### Figure S2.4.1: Design of the HLA haplotype analysis

We applied expectation-maximization algorithm to construct possible haplotypes and a logistic regression used age, sex, and the first ten principal components (PCs) to predict T1D status for covariate adjustment. We then used what we obtained to calculate haplotype score and filter reliable haplotypes through multiple testing corrections. We next computed Pearson correlation for each pair of remaining haplotypes and removed one of them if the Pearson correlation2 is greater than 0.7. Finally, we derived associations between the haplotypes remained and the AIDs and plotted the summary statistics of the associations via a heatmap.

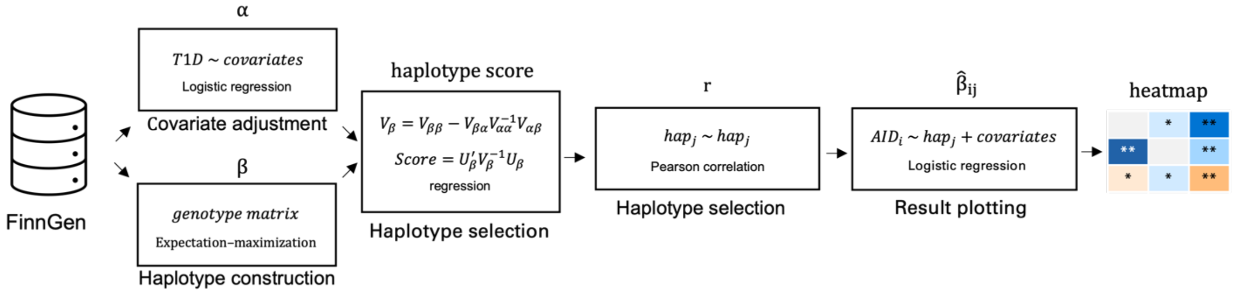

#### Figure S2.4.2: Design of the polygenic risk analysis of HLA-PGSs with 26 AIDs

Panel A shows that we extracted data from FinnGen and randomly divided all individuals into ten groups. In a 10-iteration for loop, each group was used as target set (in orange) while the remaining nine groups (in light blue) were used for model training. Panel B depicts the design of the PGS analysis. In each iteration, a weighted ridge classifier was applied on training set to derive weights ($\hat{w}_j$) for PGSs for each AID. Then we calculated PGSs for each AID using the target set and calculated partial correlation ($\rho$) between the PGSs and the AID itself for disease selection. For the diseases remained, we associated the PGSs with T1D, meta-analyzed the association statistics from the ten subgroups and plotted the results.

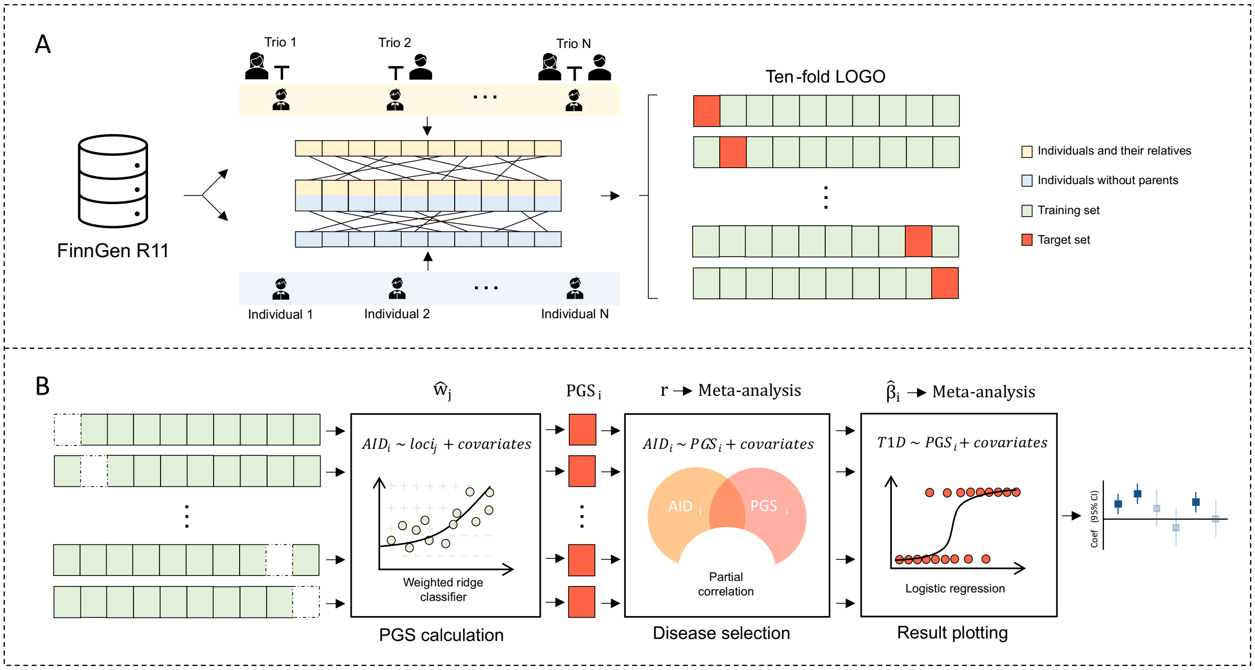


We also proposed a novel PGS method to construct PGS for T1D and other AIDs specific for HLA regions using FinnGen R11. We adopted a tenfold leave-one-group-out (LOGO) meta-analysis method (Figure S2.4.2) to avoid overfitting by making sure that the sample used for estimating the weights does not include individuals from the PGS prediction sample. We randomly divided the study population into groups.

For each AID, we used 9 groups (training set) to calculate the association between all HLA alleles and the AID with weighted ridge classifier, which can address high polymorphism and high LD in the HLA region by imposing a penalty on the size of the coefficients. The model also included age, sex, and the first ten PCs as covariates. The left-out group (validation set) was used to calculate the PGS, as a weighted sum of all imputed HLA alleles (allele dosage multiplied by its weight from the corresponding training set). After PGS normalization, we first measured partial correlation between each AID and its PGS, with the effect of the covariates removed. We dropped all the AIDs whose partial correlation with PGS for the AIDs themselves were lower than 2%. With this step, we filtered out diseases with low sample size or with weak HLA signal. Next, we conducted logistic linear regression, adjusting for age, sex, and the first ten PCs, to establish the association between PGS and T1D. The Bonferroni correction was applied to account for multiple comparisons. We repeated this process 10 times by using each different group as the validation set. The association results obtained from each of the ten validation sets were meta-analyzed using the inverse variance method.

#### 2.5 Full-PGS construction

For a given AID, we constructed both HLA PGS and non-HLA PGS separately. To understand the contribution ratio of HLA PGS and non-HLA PGS when predicting this AID, we first calculated their partial correlations ($\rho$) using a logistic regression adjusted for sex, birth year and the first 10 PCs. Then, the weight of HLA/non-HLA PGS is the contribution percentage of HLA PGS and non-HLA PGS.

$$w_{HLA\,(\boldsymbol{AID}\sim AID)} = \frac{\rho_{HLA\,(AID\sim AID)}}{\rho_{HLA\,(AID\sim AID)} + \rho_{nonHLA\,(AID\sim AID)}}$$

$$w_{nonHLA\,(\boldsymbol{AID}\sim AID)} = \frac{\rho_{nonHLA\,(AID\sim AID)}}{\rho_{HLA\,(AID\sim AID)} + \rho_{nonHLA\,(AID\sim AID)}}$$

where $\rho_{HLA\,(AID\sim AID)}$ and $\rho_{nonHLA\,(AID\sim AID)}$ are the partial correlations of HLA PGS and non-HLA PGS when we use both PGSs for an AID to predict the AID itself. Thus, the contribution ratio between HLA PGS and non-HLA PGS of this AID is $w_{HLA\,(AID\sim AID)}: w_{nonHLA\,(AID\sim AID)}$. For example, for T1D, $w_{HLA\,(T1D\sim T1D)}: w_{nonHLA\,(T1D\sim T1D)} = 0.67: 0.33$. The results as well as those of other AIDs can be found from **Table S3.5.3** column $\boldsymbol{ratio_{original}}$. Traditionally, these ratios could be used to construct full PGS for a disease to predict the disease itself.

In this study, we wondered whether we could improve PGS prediction across diseases in the full PGS context by better considering the genetic architecture (the contribution ratio of HLA PGS relative to non-HLA PGS) of our target disease, T1D. Specifically, for a given AID following pattern **1** in pTDT ($effect_{HLA} \neq 0$; $effect_{non-HLA} \neq 0$), we proposed an equation to quantify the exact contribution of HLA $PGS_{AID}$ and non-HLA $PGS_{AID}$ by taking HLA vs non-HLA contributions for both T1D and another AID into account.

$$w_{HLA\,(\boldsymbol{T1D}\sim AID)} = \frac{w_{HLA\,(T1D\sim T1D)} \times w_{HLA\,(AID\sim AID)}}{w_{HLA\,(T1D\sim T1D)} \times w_{HLA\,(AID\sim AID)} + w_{nonHLA\,(T1D\sim T1D)} \times w_{nonHLA\,(AID\sim AID)}}$$

$$w_{nonHLA\,(\boldsymbol{T1D}\sim AID)} = \frac{w_{nonHLA\,(T1D\sim T1D)} \times w_{nonHLA\,(AID\sim AID)}}{w_{HLA\,(T1D\sim T1D)} \times w_{HLA\,(AID\sim AID)} + w_{nonHLA\,(T1D\sim T1D)} \times w_{nonHLA\,(AID\sim AID)}}$$

where $w_{HLA\,(AID\sim AID)}$ and $w_{nonHLA\,(AID\sim AID)}$ are the previous weights of HLA PGS and non-HLA PGS for the AID when predicting the AID itself. $w_{HLA\,(T1D\sim T1D)}$ and $w_{nonHLA\,(T1D\sim T1D)}$ are the weights of T1D PGSs. $w_{HLA\,(T1D\sim AID)}$ and $w_{nonHLA\,(T1D\sim AID)}$ are the new weights when we use the PGSs for the AID to predict T1D.

Theoretically, with pTDT, four patterns could be defined for parental AID - offspring T1D transmissions: 1) both HLA PGS and non-HLA PGS over-/under-transmitted from unaffected parents to affected children; 2) only HLA PGS over-/under-transmitted; 3) only non-HLA PGS over-/under-transmitted; 4) neither was transmitted.

For the diseases following pattern **2** ($effect_{HLA} \neq 0$; $effect_{non-HLA} = 0$), since only HLA PGS is assumed to be over-/under-transmitted, we only used HLA PGS for these diseases to predict T1D.

$$w_{HLA\,(\boldsymbol{T1D}\sim AID)}: w_{nonHLA\,(\boldsymbol{T1D}\sim AID)} = 1: 0$$

For the diseases following pattern **3** ($effect_{HLA} = 0$; $effect_{non-HLA} \neq 0$), similarly, we only considered non-HLA PGS for these diseases to predict T1D.

$$w_{HLA\,(\boldsymbol{T1D}\sim AID)} : w_{nonHLA\,(\boldsymbol{T1D}\sim AID)} = 0:1$$

For the diseases following pattern **4** ($effect_{HLA} \neq 0$; $effect_{non-HLA} \neq 0$), neither HLA PGS nor non-HLA PGS was used for T1D prediction.

$$w_{HLA\,(\boldsymbol{T1D}\sim AID)} : w_{nonHLA\,(\boldsymbol{T1D}\sim AID)} = 0:0$$

E.g., in a scenario of cross-disease prediction using PGS for RA to predict T1D, we conducted the analyses with the following steps: 1) assigned RA to one of the four groups according to its pTDT results, 2) quantified the contribution ratio between HLA PGS and non-HLA PGS using $\rho_{HLA\,(AID\sim AID)}$, $\rho_{nonHLA\,(AID\sim AID)}$ and the proposed equations, and 3) built a Full-PGS using HLA PGS, non-HLA PGS and the ratio of their contributions. $w_{HLA\,(\boldsymbol{T1D}\sim AID)}$ and $w_{nonHLA\,(\boldsymbol{T1D}\sim AID)}$ of all the remaining AIDs can be found in **Table S3.5.3** column $\boldsymbol{ratio_{proposed}}$.

We proposed that our equation could be extended to broader scenarios for cross-disease prediction to fully utilize PGS for an exposed/correlated disease ($E$) to boost the prediction power of a target disease ($T$) under a presumed genetic architecture (e.g., partition into HLA variants vs non-HLA variants, or common variants vs rare variants). We denoted the two separate genetic components as $X$ and $Y$.

$$\boldsymbol{ratio_{proposed}} = \begin{cases} 0:0, & effect_X = 0;\ effect_Y = 0 \\ 1:0, & effect_X \neq 0;\ effect_Y = 0 \\ 0:1, & effect_X = 0;\ effect_Y \neq 0 \\ \dfrac{w_{TX} \times w_{EX}}{w_{TX} \times w_{EX} + w_{TY} \times w_{EY}} : \dfrac{w_{TY} \times w_{EY}}{w_{TX} \times w_{EX} + w_{TY} \times w_{EY}}, & effect_X \neq 0; effect_Y \neq 0 \end{cases}$$

where $w_{TX}$ and $w_{TY}$ are the weights of $X$ and $Y$ of the target disease $T$, $w_{EX}$ and $w_{EY}$ are the weight of $X$ and the weight of $Y$ of the exposed disease $E$, $\boldsymbol{ratio_{proposed}}$ is the contribution ratio between $X$ and $Y$ for $E$ when predicting $T$.

### 3. Supplementary results

#### 3.1 Basic statistics and demographic data

##### 3.1.1 Basic estimates in FinRegistry and FinnGen

Prevalence in **Table S3.1.1** stands for unadjusted life-time prevalence. The estimates of FinRegistry were calculated based on index persons whereas those of FinnGen were from the whole population in FinnGen.

**Table S3.1.1: Basic estimates from FinRegistry and FinnGen**

| | FinRegistry | | FinnGen | | FinnGen genotyped trios | |
|---|---|---|---|---|---|---|
| **Disease** | **Prevalence** | **Parental cases** | **Prevalence** | **Cases** | **Prevalence** | **Parental cases** |
| Type 1 diabetes | 0.33 % | 385 | 0.82 % | 3668 | 0.41 % | 104 |
| Adrenocortical insufficiency | 0.14 % | 166 | 0.25 % | 1100 | 0.26 % | 65 |
| Autoimmune hyperthyroidism | 0.54 % | 635 | 0.76 % | 3404 | 0.64 % | 160 |
| Autoimmune hypothyroidism | 9.85 % | 11,485 | 10.81 % | 48113 | 13.13 % | 3,299 |
| Autoimmune hemolytic anemia | 0.05 % | 55 | 0.08 % | 341 | 0.09 % | 23 |
| Allergic purpura | 0.13 % | 154 | 0.23 % | 1008 | 0.18 % | 45 |
| Vitamin B12 deficiency anaemia | 0.83 % | 962 | 0.85 % | 3782 | 1.38 % | 346 |
| Idiopathic thrombocytopenic purpura | 0.16 % | 186 | 0.21 % | 955 | 0.20 % | 50 |
| Sarcoidosis | 0.95 % | 1104 | 1.08 % | 4800 | 1.27 % | 319 |
| Primary biliary cholangitis | 0.13 % | 157 | 0.15 % | 688 | 0.21 % | 52 |
| Coeliac disease | 0.97 % | 1,129 | 1.01 % | 4497 | 1.31 % | 330 |
| Inflammatory bowel disease | 1.91 % | 2,221 | 3.08 % | 13709 | 2.75 % | 690 |
| IgA nephropathy | 0.08 % | 89 | 0.16 % | 719 | 0.09 % | 23 |
| Ankylosing spondylitis | 0.58 % | 680 | 0.77 % | 3431 | 0.74 % | 187 |
| Mixed connective tissue disease | 0.32 % | 374 | 0.48 % | 2159 | 0.51 % | 128 |
| Rheumatoid arthritis | 2.77 % | 3 224 | 3.24 % | 14426 | 4.04 % | 1 014 |
| Sjögren syndrome | 0.50 % | 585 | 0.66 % | 2938 | 0.82 % | 207 |
| Systemic sclerosis | 0.10 % | 118 | 0.17 % | 759 | 0.21 % | 54 |

| | | | | | | |
|---|---|---|---|---|---|---|
| Wegener granulomatosis | 0.07 % | 86 | 0.11 % | 475 | 0.07 % | 18 |
| Systemic lupus erythematosus | 0.18 % | 210 | 0.27 % | 1184 | 0.20 % | 51 |
| Guillain-Barre syndrome | 0.10 % | 120 | 0.11 % | 482 | 0.14 % | 34 |
| Multiple sclerosis | 0.40 % | 461 | 0.59 % | 2 606 | 0.48 % | 120 |
| Myasthenia gravis | 0.09 % | 103 | 0.11 % | 489 | 0.10 % | 25 |
| Alopecia areata | 0.15 % | 175 | 0.19 % | 856 | 0.10 % | 25 |
| Psoriasis | 2.13 % | 2 486 | 2.54 % | 11322 | 2.67 % | 671 |
| Vitiligo | 0.06 % | 65 | 0.08 % | 337 | 0.06 % | 16 |

211

##### 3.1.2 Socio-demographic characteristics in FinRegistry

To ensure that cases and the controls were comparable, we analyzed the socio-demographic variables to determine differences between the two groups. We used Cohen's D50 for ordinal variables and Chi-squared test for categorical variables. Details for the definition of socio-demographic status can be found from https://www.stat.fi/en/luokitukset/.

**Table S3.1.2: Socio-demographic characteristics of cases and controls in the matched case-control study**

| | Cases N = 14 915 | Controls N = 44 745 | Cohen's D | $\chi^2$ (P-value) |
|---|---|---|---|---|
| **Year of birth (Child): mean** | 1983.08 | 1983.02 | 0.0056 | |
| **Year of birth (Father): mean** | 1952.29 | 1952.29 | -0.0003 | |
| **Year of birth (Mother): mean** | 1954.56 | 1954.56 | -0.0003 | |
| **Number of siblings: mean** | 1.68 | 1.69 | -0.0062 | |
| **Socio-economic status: count (%)** | | | | 0.4008 (0.9997) |
| Self-employed, agriculture | 142 (1.0) | 478 (1.1) | | |
| Self-employed, non-agriculture | 647 (4.4) | 2424 (5.5) | | |
| Upper-level employees | 2293 (15.7) | 7206 (16.5) | | |
| Lower-level employees | 3865 (26.5) | 11589 (26.5) | | |
| Manual workers | 3563 (24.5) | 10905 (24.9) | | |
| Students | 1838 (12.6) | 5388 (12.3) | | |
| Pensioners | 611 (4.2) | 1303 (3) | | |
| Unknown | 1611 (11.1) | 4416 (10.1) | | |
| **Marital status: count (%)** | | | | 0.0102 (0.9196) |
| Ever married | 8835 (60.6) | 25748 (58.9) | | |
| Never married | 5736 (39.4) | 17965 (41.1) | | |
| **Number of children: mean** | 0.92 | 1.04 | -0.0875 | |
| **Occupation: count (%)** | | | | 0.3948 (1.0) |
| Armed forces | 24 (0.2) | 170 (0.4) | | |
| Managers | 300 (2.1) | 1207 (2.8) | | |
| Professionals | 2256 (15.5) | 6862 (15.7) | | |
| Technicians and associate professionals | 2169 (14.9) | 6198 (14.2) | | |
| Clerical and support workers | 773 (5.3) | 2223 (5.1) | | |
| Service and sales workers | 2776 (19.1) | 8445 (19.3) | | |
| Skilled agricultural, forestry and fishery workers | 249 (1.7) | 803 (1.8) | | |
| Craft and related trades workers | 1758 (12.1) | 4859 (11.1) | | |
| Plant and machine operators, and assemblers | 1050 (7.2) | 3775 (8.6) | | |
| Elementary occupations | 1067 (7.3) | 3179 (7.3) | | |
| Unknown | 2149 (14.7) | 5991 (13.7) | | |
| **Education level: count (%)** | | | | 0.1736 (1.0) |
| Education possibly ongoing | 6251 (42.9) | 18662 (42.7) | | |
| Upper secondary education | 3703 (25.4) | 10962 (25.1) | | |
| Specialist vocational education | 133 (0.9) | 448 (1) | | |
| Short-cycle tertiary education | 816 (5.6) | 2005 (4.6) | | |
| First-cycle higher education | 1557 (10.7) | 4679 (10.7) | | |
| Second-cycle higher education | 1268 (8.7) | 4200 (9.6) | | |
| Third-cycle higher education | 116 (0.8) | 351 (0.8) | | |
| Level of education unknown or missing | 727 (5.0) | 2405 (5.5) | | |
| **Mother tongue: count (%)** | | | | 0.2934 (0.9613) |
| Finnish | 13,746 (94.3) | 40,942 (93.7) | | |
| Sweden | 715 (4.9) | 2,146 (4.9) | | |
| Russian | 36 (0.3) | 159 (0.4) | | |
| Others | 74 (0.5) | 466 (1.1) | | |

#### 3.2 Registry-based analyses

##### 3.2.1: Matched case-control study

One of our main analyses is matched case-control study using T1D before age of 40 including 14,571 T1D cases in offspring and 43,713 controls. We reported number of cases, odds ratio (OR) with 95% confidence interval (CI) and P-values in the tables below as results.

**Table S3.2.1: Finnish population-based case-control study using T1D before age of 40 as diagnosis code**

| Disease | N (%) in case group | N (%) in control group | OR (95% CI) | P-value |
|---|---|---|---|---|
| Type 1 diabetes | 265 (0.91) | 120 (0.14) | 6.77 [5.44, 8.42] | 3.75E-66 |
| Autoimmune hypothyroidism | 3675 (12.61) | 7810 (8.93) | 1.53 [1.46, 1.60] | 4.75E-73 |
| Coeliac disease | 465 (1.6) | 664 (0.76) | 2.14 [1.90, 2.42] | 2.54E-35 |
| Rheumatoid arthritis | 1043 (3.58) | 2181 (2.49) | 1.48 [1.37, 1.60] | 1.84E-23 |
| Vitamin B12 deficiency anaemia | 350 (1.2) | 612 (0.7) | 1.76 [1.54, 2.02] | 1.40E-16 |
| Sarcoidosis | 356 (1.22) | 748 (0.86) | 1.45 [1.28, 1.65] | 1.13E-08 |
| Autoimmune hyperthyroidism | 204 (0.7) | 431 (0.49) | 1.42 [1.20, 1.68] | 4.32E-05 |
| Mixed connective tissue disease | 126 (0.43) | 248 (0.28) | 1.54 [1.24, 1.91] | 8.47E-05 |
| Sjögren syndrome | 179 (0.61) | 406 (0.46) | 1.33 [1.11, 1.59] | 1.61E-03 |
| Vitiligo | 27 (0.09) | 38 (0.04) | 2.09 [1.20, 3.63] | 1.94E-03 |
| Primary biliary cholangitis | 56 (0.19) | 101 (0.12) | 1.66 [1.20, 2.31] | 2.42E-03 |
| Autoimmune hemolytic anemia | 23 (0.08) | 32 (0.04) | 2.20 [1.28, 3.78] | 4.42E-03 |
| Myasthenia gravis | 36 (0.12) | 67 (0.08) | 1.65 [1.10, 2.48] | 1.51E-02 |
| Adrenocortical insufficiency | 53 (0.18) | 113 (0.13) | 1.42 [1.02, 1.97] | 3.51E-02 |
| Psoriasis | 665 (2.28) | 1821 (2.08) | 1.10 [1.01, 1.21] | 3.53E-02 |
| Systemic lupus erythematosus | 63 (0.22) | 147 (0.17) | 1.29 [0.96, 1.73] | 9.18E-02 |
| IgA nephropathy | 28 (0.1) | 61 (0.07) | 1.40 [0.89, 2.18] | 1.43E-01 |
| Systemic sclerosis | 36 (0.12) | 82 (0.09) | 1.33 [0.90, 1.97] | 1.55E-01 |
| Alopecia areata | 51 (0.18) | 124 (0.14) | 1.24 [0.89, 1.72] | 1.99E-01 |
| Guillain-Barre syndrome | 35 (0.12) | 85 (0.1) | 1.25 [0.84, 1.86] | 2.76E-01 |
| Allergic purpura | 44 (0.15) | 110 (0.13) | 1.20 [0.85, 1.71] | 2.98E-01 |
| Inflammatory bowel disease | 573 (1.97) | 1648 (1.89) | 1.05 [0.95, 1.16] | 3.19E-01 |
| Wegener granulomatosis | 25 (0.09) | 61 (0.07) | 1.24 [0.78, 1.98] | 3.66E-01 |
| Ankylosing spondylitis | 161 (0.55) | 519 (0.59) | 0.93 [0.78, 1.11] | 4.11E-01 |
| Idiopathic thrombocytopenic purpura | 50 (0.17) | 136 (0.16) | 1.10 [0.80, 1.53] | 5.50E-01 |
| Multiple sclerosis | 111 (0.38) | 350 (0.4) | 0.94 [0.76, 1.17] | 5.91E-01 |

#### Figure S3.2.2: Association between parental AIDs and T1D in offspring by sex

Panel A shows how T1D among children is associated with paternal AIDs and maternal AIDs. Panel B is the associations stratified by sex of children. Only parental AIDs significantly associated with T1D (**Fig.2**) are displayed.

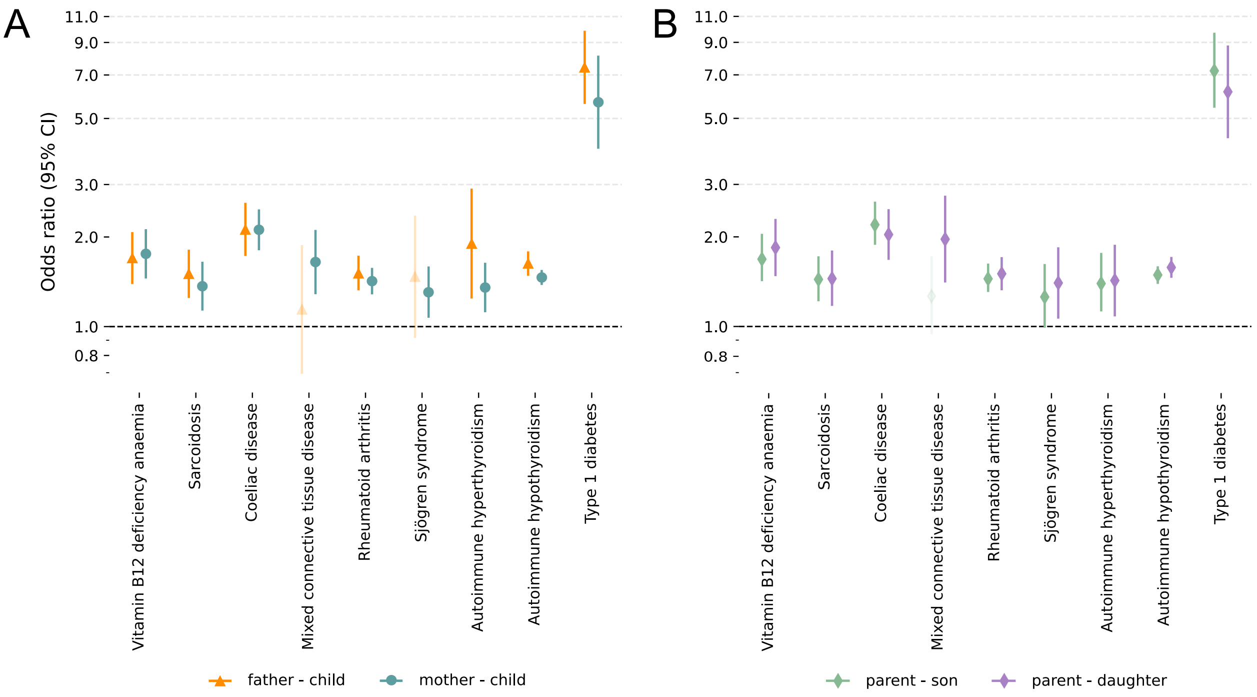


#### 3.2.3 Association between parental AIDs and age of T1D onset in offspring by sex

We then assessed how parental AIDs affected age at onset of T1D in offspring. We excluded individuals without onset age. The median age at diagnosis (the first record of the ICD-code) of T1D was 12.7 years (IQR=[8.0,20.0]) with men (13.6 years, IQR=[8.6,21.1]) being older than women (11.6 years, IQR=[7.3,18.2]). In a multivariate regression analysis adjusted by all the matching factors, only parental T1D were significantly ($P<0.05/9$) associated with the age of T1D onset in offspring (Table S3.2.3.1). The observed onset age among individuals with T1D was 1.57 [0.54-2.60] (P =0.0028) years younger if at least one parent had T1D. We further observed that this association mainly came from the association between paternal T1D and T1D onset among sons while mothers with T1D had no impact on the T1D onset of their offspring (**Table S3.2.3.2**). To test if these associations could be biased by potential data structure due to e.g., average age of onset for different diseases, the change of ICD definitions and gradually improved healthcare policies over the years, we split the data by children's birthyear by median of their birthyear (1960-1979 and 1980-1999). We conducted the same analysis and obtained consistent results (**Table S3.2.3.3**).

**Table 3.2.3.1: Association between the age of T1D onset in offspring and parental AID status**

| Parental AID | # AID cases | log(OR) [95% CI] | P |
|---|---|---|---|
| Type 1 diabetes | 254 | -1.57 [-2.6, -0.54] | 0.0028 |
| Autoimmune hypothyroidism | 3 469 | -0.06 [-0.37, 0.26] | 0.7264 |
| Coeliac disease | 458 | -0.8 [-1.57, -0.04] | 0.0399 |
| Rheumatoid arthritis | 1 017 | -0.55 [-1.08, -0.03] | 0.0397 |
| Sarcoidosis | 333 | -0.32 [-1.22, 0.57] | 0.4803 |
| Vitamin B12 deficiency anemia | 357 | 0.88 [0, 1.76] | 0.0502 |
| Autoimmune hyperthyroidism | 200 | -1.15 [-2.31, 0] | 0.0495 |
| Mixed connective tissue disease | 126 | -1.02 [-2.46, 0.42] | 0.1666 |
| Sjögren syndrome | 176 | 0.28 [-0.94, 1.51] | 0.6483 |

**Table 3.2.3.2: Relationship between the age of T1D onset in offspring and parental T1D status stratified by sex of parents and offspring**

| Relationship | # Parental T1D cases | log(OR) [95% CI] | P |
|---|---|---|---|
| father – daughter | 60 | -1.88 [-3.92, 0.16] | 0.0704 |
| mother – daughter | 34 | -0.4 [-3.1, 2.3] | 0.7727 |
| father – son | 113 | -1.85 [-3.41, -0.29] | 0.0201 |
| mother – son | 49 | -1.17 [-3.53, 1.18] | 0.3277 |

**Table 3.2.3.3: Relationship between the age of T1D onset in offspring and parental T1D status stratified by sex of parents and offspring with the offspring's birthyear within 1980-1999**

| Relationship | # Parental T1D cases | log(OR) [95% CI] | P |
|---|---|---|---|
| parent – child | 242 | -1.45 [-2.35, -0.54] | 0.0017 |
| father – daughter | 56 | -1.59 [-3.41, 0.22] | 0.0851 |
| mother – daughter | 33 | -0.03 [-2.38, 2.32] | 0.9832 |
| father – son | 109 | -1.76 [-3.13, -0.39] | 0.0118 |
| mother – son | 47 | -1.42 [-3.49, 0.64] | 0.1774 |

##### 3.2.4: Population-based sensitivity analyses

##### Table S3.2.4.1: Matched case-control study using T1D before age of 20 as diagnosis code

| Disease | N (%) in case group | N (%) in control group | OR (95% CI) | P-value |
|---|---|---|---|---|
| Type 1 diabetes | 171 (0.84) | 48 (0.08) | 10.9 [7.90, 15.04] | 6.81E-48 |
| Autoimmune hypothyroidism | 2396 (11.79) | 5014 (8.24) | 1.55 [1.46, 1.64] | 2.31E-52 |
| Coeliac disease | 345 (1.7) | 485 (0.8) | 2.18 [1.90, 2.51] | 1.39E-27 |
| Rheumatoid arthritis | 622 (3.06) | 1276 (2.1) | 1.50 [1.36, 1.66] | 1.78E-15 |
| Sarcoidosis | 168 (0.83) | 337 (0.55) | 1.48 [1.27, 1.73] | 7.65E-07 |
| Vitamin B12 deficiency anaemia | 244 (1.2) | 500 (0.82) | 1.50 [1.25, 1.81] | 2.08E-05 |
| Autoimmune hyperthyroidism | 155 (0.76) | 330 (0.54) | 1.41 [1.16, 1.71] | 4.57E-04 |
| Mixed connective tissue disease | 88 (0.43) | 168 (0.28) | 1.59 [1.22, 2.06] | 4.98E-04 |
| Systemic lupus erythematosus | 101 (0.5) | 281 (0.46) | 1.77 [1.25, 2.51] | 1.41E-03 |
| Primary biliary cholangitis | 23 (0.11) | 39 (0.06) | 1.79 [1.17, 2.73] | 6.75E-03 |
| Vitiligo | 35 (0.17) | 59 (0.1) | 1.77 [1.05, 2.96] | 3.06E-02 |
| Alopecia areata | 8 (0.04) | 20 (0.03) | 1.47 [1.00, 2.17] | 4.94E-02 |
| Psoriasis | 21 (0.1) | 52 (0.09) | 1.09 [0.98, 1.22] | 1.18E-01 |
| Adrenocortical insufficiency | 38 (0.19) | 85 (0.14) | 1.35 [0.92, 1.98] | 1.25E-01 |
| IgA nephropathy | 448 (2.2) | 1237 (2.03) | 1.43 [0.85, 2.41] | 1.74E-01 |
| Idiopathic thrombocytopenic purpura | 50 (0.25) | 86 (0.14) | 1.32 [0.88, 1.97] | 1.82E-01 |
| Ankylosing spondylitis | 21 (0.1) | 44 (0.07) | 0.87 [0.70, 1.09] | 2.21E-01 |
| Systemic sclerosis | 25 (0.12) | 57 (0.09) | 1.33 [0.83, 2.12] | 2.37E-01 |
| Inflammatory bowel disease | 38 (0.19) | 78 (0.13) | 1.05 [0.94, 1.18] | 3.57E-01 |
| Wegener granulomatosis | 25 (0.12) | 65 (0.11) | 1.33 [0.69, 2.57] | 3.96E-01 |
| Myasthenia gravis | 31 (0.15) | 80 (0.13) | 1.21 [0.73, 2.02] | 4.61E-01 |
| Allergic purpura | 432 (2.12) | 1239 (2.04) | 1.16 [0.77, 1.76] | 4.71E-01 |
| Sjögren syndrome | 13 (0.06) | 38 (0.06) | 1.08 [0.86, 1.36] | 5.10E-01 |
| Guillain-Barre syndrome | 104 (0.51) | 361 (0.59) | 1.16 [0.73, 1.84] | 5.35E-01 |
| Autoimmune hemolytic anemia | 34 (0.17) | 78 (0.13) | 1.20 [0.53, 2.73] | 6.58E-01 |
| Multiple sclerosis | 89 (0.44) | 275 (0.45) | 0.98 [0.77, 1.25] | 8.79E-01 |

#### Table S3.2.4.2: Association between parental AIDs and T1D in children by sex

| Disease | Parent | OR (95% CI) | P-value | Child | OR (95% CI) | P-value |
|---|---|---|---|---|---|---|
| Type 1 diabetes | Father | 12.43 [8.25, 18.72] | 1.76E-33 | Son | 13.27 [8.48, 20.78] | 1.15E-29 |
| | Mother | 8.5 [5.06, 14.27] | 6.33E-16 | Daughter | 8.75 [5.48, 13.99] | 1.19E-19 |
| Autoimmune hyperthyroidism | Father | 1.41 [0.89, 2.25] | 1.46E-01 | Son | 1.6 [1.25, 2.06] | 2.13E-04 |
| | Mother | 1.42 [1.15, 1.75] | 1.13E-03 | Daughter | 1.18 [0.87, 1.61] | 2.86E-01 |
| Autoimmune hypothyroidism | Father | 1.62 [1.46, 1.81] | 5.62E-19 | Son | 1.49 [1.38, 1.6] | 5.08E-26 |
| | Mother | 1.49 [1.4, 1.58] | 1.16E-37 | Daughter | 1.64 [1.5, 1.79] | 1.16E-28 |
| Vitamin B12 deficiency anaemia | Father | 1.62 [1.22, 2.15] | 7.83E-04 | Son | 1.46 [1.13, 1.89] | 3.39E-03 |
| | Mother | 1.43 [1.11, 1.83] | 5.16E-03 | Daughter | 1.55 [1.18, 2.05] | 1.92E-03 |
| Sarcoidosis | Father | 1.64 [1.33, 2.01] | 3.43E-06 | Son | 1.36 [1.11, 1.66] | 3.42E-03 |
| | Mother | 1.3 [1.03, 1.64] | 2.68E-02 | Daughter | 1.68 [1.32, 2.14] | 2.39E-05 |
| Coeliac disease | Father | 2.01 [1.59, 2.53] | 3.33E-09 | Son | 2.29 [1.9, 2.76] | 2.77E-18 |
| | Mother | 2.26 [1.9, 2.69] | 7.95E-20 | Daughter | 2.05 [1.65, 2.54] | 5.55E-11 |
| Mixed connective tissue disease | Father | 1.69 [0.96, 3] | 7.11E-02 | Son | 1.3 [0.92, 1.84] | 1.44E-01 |
| | Mother | 1.56 [1.17, 2.08] | 2.70E-03 | Daughter | 2.09 [1.4, 3.11] | 2.79E-04 |
| Rheumatoid arthritis | Father | 1.4 [1.19, 1.66] | 6.88E-05 | Son | 1.39 [1.21, 1.58] | 1.29E-06 |
| | Mother | 1.53 [1.35, 1.72] | 6.24E-12 | Daughter | 1.67 [1.43, 1.94] | 4.50E-11 |
| Systemic lupus erythematosus | Father | 1.06 [0.47, 2.4] | 8.96E-01 | Son | 1.46 [0.89, 2.38] | 1.32E-01 |
| | Mother | 1.98 [1.34, 2.92] | 6.30E-04 | Daughter | 2.18 [1.31, 3.6] | 2.52E-03 |

#### 3.3 Shared genetic background in population

Given that for T1D, the strongest genetic associations are for Class II haplotypes, in particular HLA-DR3 and DR4 haplotypes, we constructed 19 *DRB1-DQA1-DQB1* haplotypes based on 64 imputed alleles of these genes in FinnGen. We examined 19 haplotypes (frequencies > 0.5%, **Table S3.3.1.1**), of which 15 were strongly associated with T1D ($P<1.0 \times 10^{-4}$), with *DRB1*04:01-DQA1*03:01-DQB1*03:02* (OR=7.54 [7.00, 8.13], $P<1.0 \times 10^{-602}$) conferring the strongest susceptibility and *DRB1*15:01-DQA1*01:02-DQB1*06:02* (0.06 [0.04, 0.07], $P=2.0\times10^{-106}$) the strongest protection. We then tested the association of these 15 haplotypes with the 25 other included AIDs using logistic regression and considering the age, sex, and first ten principal components (PCs) as covariates (**Figure S3.3.1.2**). Many of these T1D-associated haplotypes were also associated with other AIDs ($P<1.0 \times10^{-4}$). For example, the T1D risk haplotype *DRB1*03:01-DQA1*05:01-DQB1*02:01* increased the risk of both CD (15.83 [14.56, 17.20], $P<1.0 \times10^{-918}$) and autoimmune hyperthyroidism (2.42 [2.23, 2.63], $P=6.3\times10^{-100}$), and *DRB1*04:01-DQA1*03:01-DQB1*03:02* increased the risk of RA (1.93 [1.84, 2.02], $P=5.1 \times10^{-174}$). Also, opposite effects were seen for some AIDs: the strongest haplotype protecting against T1D (*DRB1*15:01-DQA1*01:02-DQB1*06:02*) was the lead risk haplotype for MS (2.85 [2.60, 3.12], $P=7.9\times10^{-112}$) while the strongest T1D susceptibility haplotype (*DRB1*04:01-DQA1*03:01-DQB1*03:02*) protected against IBD (0.85 [0.80, 0.90], $P=4.7\times10^{-8}$).

##### 3.3.1 Analyses in HLA regions – haplotype analysis

##### Table S3.3.1.1: Summary statistics of haplotype-specific score tests for T1D

| Haplotype | Frequency | Haplotype score | P-value $_{score}$ |
|---|---|---|---|
| DRB1*15:01-DQA1*01:02-DQB1*06:02 | 0.14112373 | -29.639428 | 1.00E-193 |
| DRB1*13:01-DQA1*01:03-DQB1*06:03 | 0.08985561 | -16.891367 | 1.00E-64 |
| DRB1*01:01-DQA1*01:01-DQB1*05:01 | 0.18825249 | -15.024028 | 1.00E-51 |
| DRB1*11:01-DQA1*05:05-DQB1*03:01 | 0.0362712 | -13.010508 | 1.00E-38 |
| DRB1*07:01-DQA1*02:01-DQB1*03:03 | 0.01537059 | -9.2391112 | 1.00E-20 |
| DRB1*12:01-DQA1*05:05-DQB1*03:01 | 0.02637552 | -7.8918718 | 3.00E-15 |
| DRB1*07:01-DQA1*02:01-DQB1*02:02 | 0.04085991 | -6.1855557 | 6.19E-10 |
| DRB1*10:01-DQA1*01:05-DQB1*05:01 | 0.00854685 | -5.8481741 | 4.97E-09 |
| DRB1*14:54-DQA1*01:04-DQB1*05:03 | 0.00615272 | -5.7002677 | 1.20E-08 |
| DRB1*04:08-DQA1*03:03-DQB1*03:01 | 0.01166135 | -5.2422534 | 1.59E-07 |
| DRB1*13:01-DQA1*03:02-DQB1*03:03 | 0.00685471 | -5.1770243 | 2.25E-07 |
| DRB1*08:01-DQA1*04:01-DQB1*04:02 | 0.09734738 | -3.0819368 | 2.05E-03 |
| DRB1*16:01-DQA1*01:02-DQB1*05:02 | 0.00813475 | -2.4329221 | 1.50E-02 |
| DRB1*04:01-DQA1*03:03-DQB1*03:01 | 0.01094099 | -2.3841454 | 1.71E-02 |
| DRB1*09:01-DQA1*03:02-DQB1*03:03 | 0.03477909 | -1.2612753 | 2.07E-01 |
| DRB1*13:02-DQA1*01:02-DQB1*06:04 | 0.03329611 | 4.75611941 | 1.97E-06 |
| DRB1*04:04-DQA1*03:01-DQB1*03:02 | 0.04202424 | 18.4916972 | 1.00E-76 |
| DRB1*03:01-DQA1*05:01-DQB1*02:01 | 0.10353824 | 26.2203348 | 1.00E-151 |
| DRB1*04:01-DQA1*03:01-DQB1*03:02 | 0.07418242 | 62.7742651 | 1.00E-858 |

**Figure S3.3.1.2: HLA haplotypes strongly associated with T1D and their associations with other AIDs in FinnGen (3,668 T1D cases and 436,149 T1D controls)**

Based on **S3.3.1.1**, we extended the analysis to other AIDs. The strength of the association is shown as a heatmap with blue depicting a susceptible haplotype associated with the AID in question. The haplotypes are sorted by the P values for T1D as shown on the left.

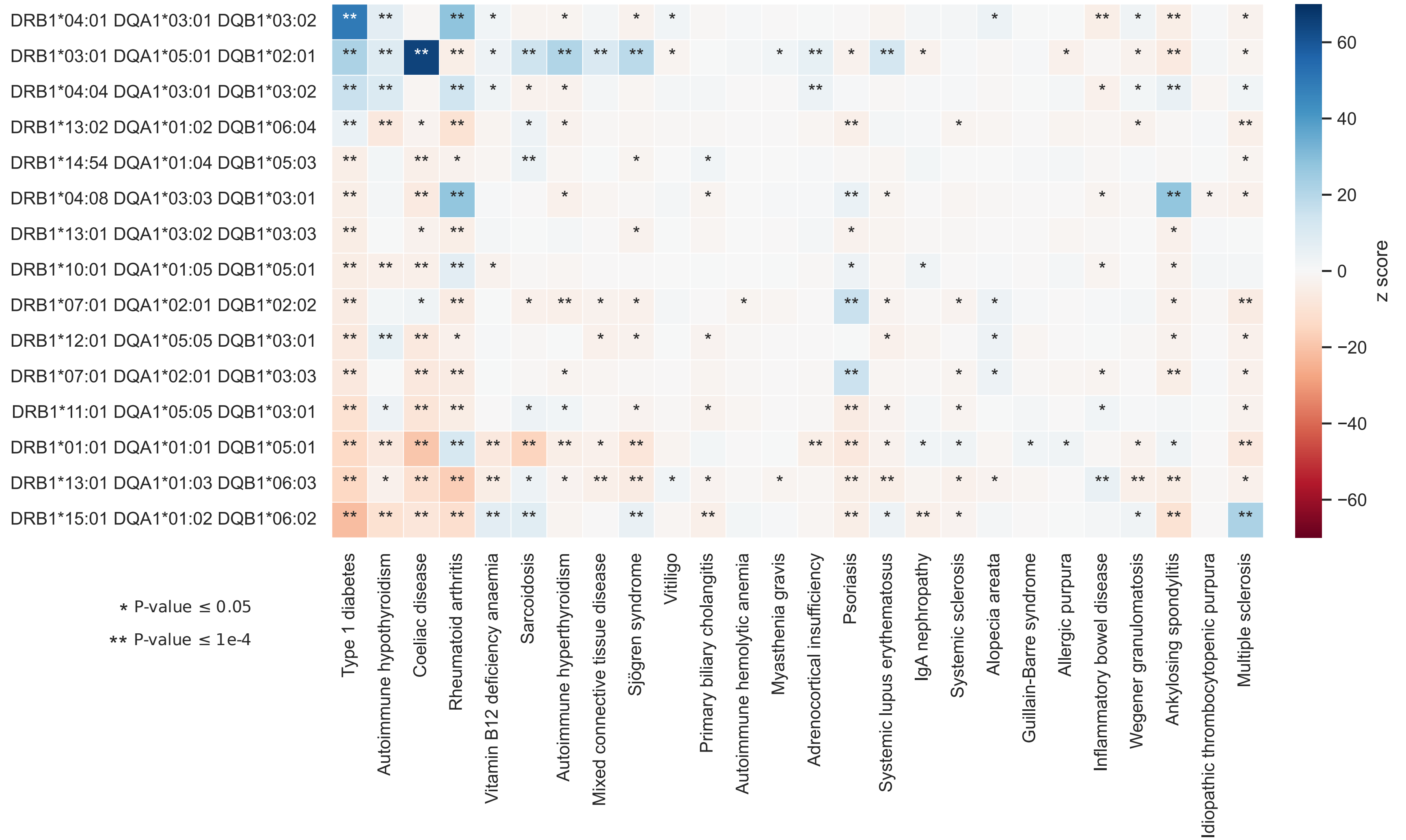

##### 3.3.2 Analyses in HLA regions – amino acid analysis

The results of the HLA amino acid analyses that showed significant association are presented in Table **S3.3.2.1** ($P<1.0 \times 10^{-4}$). The results further showed the complexity in the HLA regions. While T1D and some AIDs presented significant associations with amino acid with effects all in same direction, we also observed antagonistic HLA pleiotropy among some AIDs. For example, four residues (Ala, Arg, Glu, and Lys) at the amino acid position 72 of the *HLA-DRB1* gene exerted significant positive associations for risk of T1D and CD (Figure S3.3.2.2). However, three of them (Lys72, Glu72, and Ala72) displayed opposite effects between T1D and other AIDs, including MS and IBD.

**Table S3.3.2.1: Summary statistics of associations between amino acid positions and AIDs**

See excel file

**Figure S3.3.2.2: Associations of HLA-DRB1 amino acid positions 13 and 72 with risk for each of the AIDs**

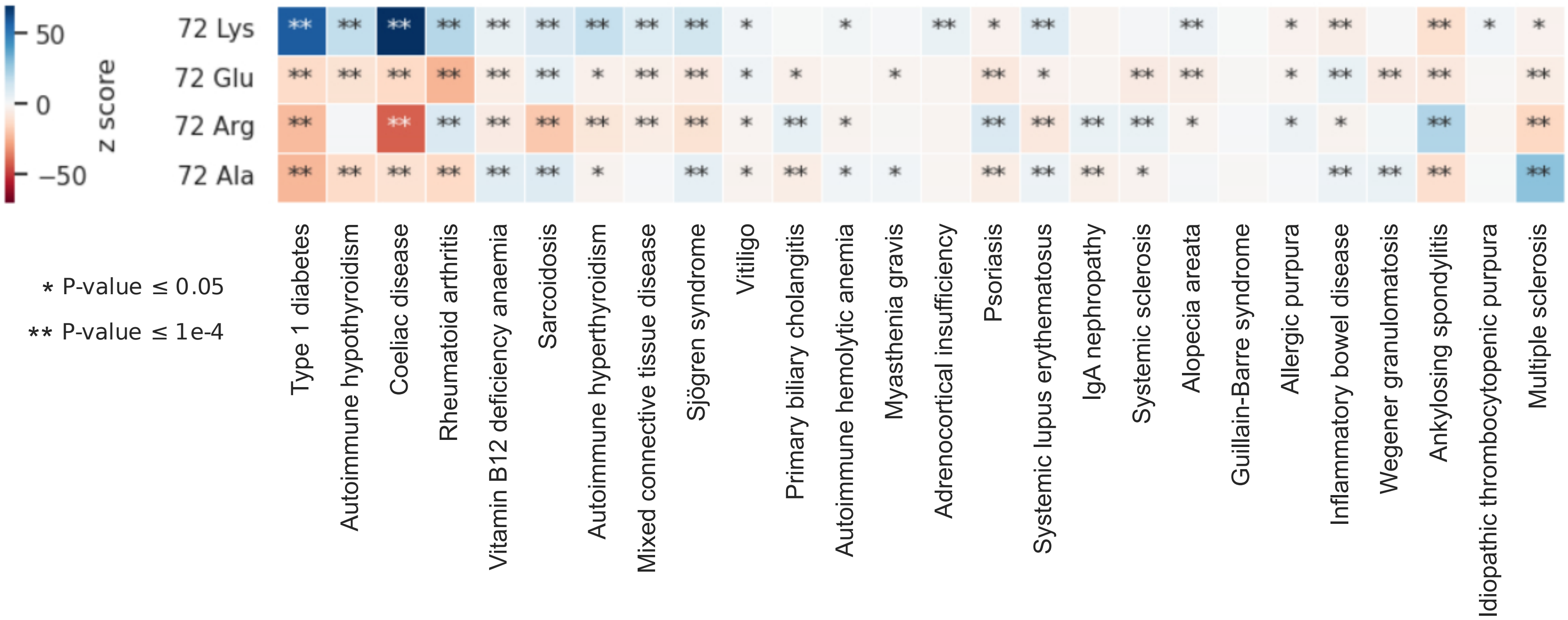

##### 3.3.3 Analyses in HLA regions – PGS analysis

##### Table S3.3.3.1: Weights of HLA alleles for constructing HLA PGS

See excel file

##### Table S3.3.3.2: Associations between HLA PGS and AIDs

This table includes number of AID cases (N), number of co-occurrences of the given AID among the cases (N.co), odds ratio (OR) with 95% CI, and partial correlation (ρ) between AIDs and HLA-PGS for the AIDs. We computed ρ to filter the AIDs (|ρ|>2%) that would be used for analysis of association between HLA PGS and T1D.

| | Number of cases | | Association between an AID and HLA polygenic risk for this AID | | | Association between T1D and HLA polygenic risk for this AID | | |
|---|---|---|---|---|---|---|---|---|
| Disease | N | N.co | OR [95% CI] | P-value | ρ(%) | OR [95% CI] | P-value | ρ(%) |
| Type 1 diabetes | 1,182 | 1 182 | 5.33 [4.79, 5.93] | 3.74E-207 | 29.98 | 5.33 [4.79, 5.93] | 3.74E-207 | 29.98 |
| Autoimmune hypothyroidism | 2,580 | 179 | 1.23 [1.18, 1.28] | 1.90E-22 | 6.39 | 1.55 [1.44, 1.67] | 6.43E-31 | 9.66 |
| Coeliac disease | 292 | 62 | 2.79 [2.55, 3.06] | 1.28E-107 | 17.07 | 2.07 [1.94, 2.22] | 2.39E-96 | 17.06 |
| Rheumatoid arthritis | 712 | 29 | 1.63 [1.52, 1.76] | 7.14E-41 | 8.92 | 2.10 [1.95, 2.27] | 1.79E-78 | 15.06 |
| Vitamin B12 deficiency anaemia | 195 | 5 | 1.18 [1.02, 1.36] | 2.81E-02 | 1.49 | | | |
| Sarcoidosis | 273 | 8 | 1.47 [1.29, 1.66] | 1.64E-09 | 3.97 | 1.15 [1.07, 1.24] | 2.16E-04 | 2.39 |
| Autoimmune hyperthyroidism | 161 | 14 | 1.61 [1.41, 1.85] | 1.99E-12 | 4.68 | 1.19 [1.11, 1.27] | 7.18E-07 | 3.49 |
| Mixed connective tissue disease | 102 | <5 | 1.17 [0.97, 1.41] | 1.04E-01 | 1.05 | | | |
| Sjögren syndrome | 154 | <5 | 1.39 [1.20, 1.60] | 9.30E-06 | 2.93 | 1.29 [1.21, 1.39] | 3.15E-13 | 5.21 |
| Vitiligo | 15 | <5 | 2.21 [1.32, 3.68] | 2.39E-03 | 2.04 | 1.83 [1.69, 1.97] | 3.39E-53 | 17.43 |
| Primary biliary cholangitis | 38 | <5 | 1.71 [1.29, 2.28] | 2.11E-04 | 2.43 | 1.43 [1.33, 1.54] | 4.39E-21 | 7.06 |
| Autoimmune hemolytic anemia | 17 | 0 | 0.97 [0.6, 1.55] | 8.85E-01 | -0.11 | | | |
| Myasthenia gravis | 19 | 0 | 1.06 [0.68, 1.66] | 7.82E-01 | 0.17 | | | |
| Adrenocortical insufficiency | 67 | 6 | 1.59 [1.27, 1.99] | 5.07E-05 | 2.67 | 2.30 [2.13, 2.49] | 4.17E-100 | 17.43 |
| Psoriasis | 547 | 22 | 1.51 [1.40, 1.62] | 6.93E-30 | 7.60 | 1.11 [1.04, 1.20] | 3.57E-03 | 2.40 |
| Systemic lupus erythematosus | 53 | <5 | 1.77 [1.39, 2.25] | 4.40E-06 | 3.00 | 1.40 [1.31, 1.51] | 6.24E-21 | 6.44 |
| IgA nephropathy | 24 | 0 | 1.06 [0.71, 1.57] | 7.83E-01 | 0.17 | | | |

| | | | | | | | | |
|---|---|---|---|---|---|---|---|---|
| Systemic sclerosis | 41 | 0 | 1.07 [0.79, 1.45] | 6.44E-01 | 0.32 | | | |
| Alopecia areata | 47 | 6 | 1.33 [1.00, 1.77] | 5.07E-02 | 1.29 | | | |
| Guillain-Barre syndrome | 33 | <5 | 1.09 [0.78, 1.53] | 6.02E-01 | 0.34 | | | |
| Allergic purpura | 53 | 6 | 1.20 [0.92, 1.56] | 1.86E-01 | 0.87 | | | |
| Inflammatory bowel disease | 670 | 18 | 1.29 [1.19, 1.39] | 2.04E-10 | 4.19 | 0.64 [0.60, 0.69] | 1.15E-30 | -9.10 |
| Wegener granulomatosis | 23 | 0 | 1.37 [0.90, 2.11] | 1.45E-01 | 0.99 | | | |
| Ankylosing spondylitis | 187 | <5 | 2.22 [2.03, 2.42] | 5.92E-72 | 14.08 | 0.93 [0.86, 1.00] | 6.13E-02 | -1.79 |
| Idiopathic thrombocytopenic purpura | 46 | <5 | 1.11 [0.84, 1.46] | 4.77E-01 | 0.46 | | | |
| Multiple sclerosis | 128 | 8 | 1.95 [1.67, 2.29] | 1.11E-16 | 5.52 | 0.81 [0.75, 0.88] | 2.40E-07 | -4.16 |

#### Table S3.3.3.3: Associations between HLA PGS and AIDs using leave-one-group-out method

To fully utilize the data, we conducted a sensitivity analysis using LOGO method (**Figure S2.4.2**) to conduct HLA PGS analysis. The results (**Table S3.3.2.2**) are very similar to those from the core analysis (**Table S3.3.2.1**).

| | Number of cases | | Association between an AID and HLA polygenic risk for this AID | | | Association between T1D and HLA polygenic risk for this AID | | |
|---|---|---|---|---|---|---|---|---|
| **Disease** | **N** | **N.co** | **OR (95% CI)** | **P-value** | **ρ(%)** | **OR (95% CI)** | **P-value** | **ρ(%)** |
| Type 1 diabetes | 3 307 | 3 307 | 3.99 [3.84, 4.16] | 5.27E-94 | 13.61 | 3.99 [3.84, 4.16] | 5.27E-94 | 13.61 |
| Autoimmune hypothyroidism | 41 367 | 553 | 1.25 [1.23, 1.27] | 3.22E-26 | 6.74 | 1.63 [1.58, 1.69] | 6.81E-16 | 4.95 |
| Coeliac disease | 3 792 | 159 | 2.72 [2.66, 2.79] | 9.47E-127 | 15.78 | 1.81 [1.75, 1.87] | 8.85E-22 | 7.89 |
| Rheumatoid arthritis | 12 230 | 82 | 1.64 [1.61, 1.67] | 6.65E-58 | 9.17 | 1.91 [1.85, 1.98] | 7.21E-28 | 6.92 |
| Vitamin B12 deficiency anaemia | 3132 | 30 | 1.31 [1.23, 1.39] | 4.40E-03 | 2.28 | 1.57 [1.51, 1.63] | 3.96E-10 | 3.84 |
| Sarcoidosis | 4117 | 34 | 1.55 [1.5, 1.6] | 6.75E-14 | 4.33 | 1.11 [1.07, 1.16] | 1.79E-01 | 0.95 |
| Autoimmune hyperthyroidism | 3 054 | 48 | 1.51 [1.46, 1.55] | 1.30E-14 | 4.33 | 1.27 [1.22, 1.33] | 8.27E-04 | 2.63 |
| Mixed connective tissue disease | 1898 | 13 | 1.31 [1.26, 1.37] | 1.12E-03 | 2.09 | 1.65 [1.59, 1.72] | 3.85E-20 | 5.73 |
| Sjögren syndrome | 2479 | 7 | 1.52 [1.47, 1.59] | 6.16E-10 | 3.91 | 1.22 [1.16, 1.28] | 4.79E-02 | 2.09 |
| Vitiligo | 305 | 6 | 1.54 [1.38, 1.72] | 4.75E-02 | 1.28 | | | |

| | | | | | | | | |
|---|---|---|---|---|---|---|---|---|
| Primary biliary cholangitis | 574 | <5 | 1.62 [1.47, 1.78] | 7.21E-03 | 2.00 | 1.38 [1.31, 1.44] | 3.32E-06 | 3.18 |
| Autoimmune hemolytic anemia | 280 | <5 | 1.23 [1.09, 1.39] | 4.37E-01 | 0.53 | | | |
| Myasthenia gravis | 428 | 0 | 1.16 [1.06, 1.33] | 4.25E-01 | 0.51 | | | |
| Adrenocortical insufficiency | 947 | 20 | 1.25 [1.15, 1.36] | 1.50E-01 | 1.13 | | | |
| Psoriasis | 9 900 | 67 | 1.49 [1.46, 1.51] | 7.30E-41 | 7.51 | 1.1 [1.06, 1.14] | 1.47E-01 | 0.94 |
| Systemic lupus erythematosus | 1040 | 6 | 1.44 [1.35, 1.53] | 3.80E-03 | 2.12 | 1.33 [1.27, 1.39] | 7.14E-04 | 2.92 |
| IgA nephropathy | 647 | <5 | 1.32 [1.21, 1.43] | 1.23E-01 | 1.12 | | | |
| Systemic sclerosis | 641 | <5 | 1.58 [1.45, 1.72] | 1.90E-02 | 1.81 | | | |
| Alopecia areata | 755 | 15 | 1.33 [1.23, 1.45] | 8.01E-02 | 1.26 | | | |
| Guillain-Barre syndrome | 418 | <5 | 1.08 [0.98, 1.19] | 5.15E-01 | 0.25 | | | |
| Allergic purpura | 893 | 10 | 1.24 [1.14, 1.36] | 2.36E-01 | 1.05 | | | |
| Inflammatory bowel disease | 12210 | 69 | 1.18 [1.16, 1.2] | 4.48E-06 | 2.90 | 0.66 [0.63, 0.68] | 1.82E-11 | -4.08 |
| Wegener granulomatosis | 418 | 0 | 1.93 [1.67, 2.24] | 1.83E-02 | 1.86 | | | |
| Ankylosing spondylitis | 3062 | 18 | 2.34 [2.29, 2.39] | 5.43E-117 | 15.22 | 0.98 [0.94, 1.01] | 4.83E-01 | -0.27 |
| Idiopathic thrombocytopenic purpura | 795 | 6 | 1.06 [0.97, 1.15] | 4.06E-01 | 0.24 | | | |
| Multiple sclerosis | 2396 | 18 | 1.81 [1.74, 1.88] | 5.74E-21 | 5.16 | 0.78 [0.75, 0.82] | 3.32E-03 | -2.16 |

#### Table S3.3.3.4: Model setting for ridge classifier, lasso regression and elastic net

We compared all three models, ridge classifier, lasso regression, and elastic net, for HLA PGS construction. For each model, we used grid search to find the best parameters in the given range, with the range chosen based on the computation time.

| Model | Parameter range |
|---|---|
| Ridge classifier | Alpha: 1-10000 |
| Lasso regression | C: 0.001-1000 |
| Elastic Net | C: 0.001-1000; l1:l2 ratio: 0.1-0.9 |

#### Table S3.3.3.5: Comparison among ridge classifier, lasso regression and elastic net

We found that overall ridge regression outperformed the other two in our analysis regarding the increment of pseudo-$R^2$ of the model and AUC when we use HLA-PGS built from each model for the AIDs to predict the incidence risk of the same AIDs. Therefore, we chose to use ridge regression in our main analyses.

| Disease | Ridge classifier | | Lasso regression | | Elastic net | |
|---|---|---|---|---|---|---|
| | Δpseudo-$R^2$ | ΔAUC | Δpseudo-$R^2$ | ΔAUC | Δpseudo-$R^2$ | ΔAUC |
| Type 1 diabetes | 0.1615<br>SD=0.0034 | 0.0322<br>SD=0.0029 | 0.1220<br>SD=0.0028 | 0.0259<br>SD=0.0023 | 0.1220<br>SD=0.0028 | 0.0258<br>SD=0.0023 |
| Autoimmune hypothyroidism | 0.0058<br>SD=0.0008 | 0.0084<br>SD=0.0026 | 0.0051<br>SD=0.0008 | 0.0072<br>SD=0.0026 | 0.0051<br>SD=0.0008 | 0.0072<br>SD=0.0026 |
| Coeliac disease | 0.1564<br>SD=0.0055 | 0.2511<br>SD=0.0183 | 0.1380<br>SD=0.0056 | 0.2292<br>SD=0.0191 | 0.1380<br>SD=0.0056 | 0.2292<br>SD=0.0191 |
| Rheumatoid arthritis | 0.0279<br>SD=0.0022 | 0.0521<br>SD=0.0098 | 0.0205<br>SD=0.0018 | 0.0397<br>SD=0.0085 | 0.0205<br>SD=0.0018 | 0.0397<br>SD=0.0085 |
| Sarcoidosis | 0.0128<br>SD=0.0019 | 0.0406<br>SD=0.0134 | 0.0085<br>SD=0.0018 | 0.0291<br>SD=0.0121 | 0.0085<br>SD=0.0018 | 0.0292<br>SD=0.0123 |
| Vitamin B12 deficiency anaemia | 0.0029<br>SD=0.0014 | 0.0013<br>SD=0.0045 | 0.0021<br>SD=0.0013 | 0.0011<br>SD=0.0045 | 0.0021<br>SD=0.0013 | 0.0011<br>SD=0.0045 |
| Autoimmune hyperthyroidism | 0.0235<br>SD=0.0048 | 0.0610<br>SD=0.0163 | 0.0185<br>SD=0.0043 | 0.0449<br>SD=0.0161 | 0.0185<br>SD=0.0043 | 0.0449<br>SD=0.0161 |
| Mixed connective tissue disease | 0.0025<br>SD=0.0012 | 0.0076<br>SD=0.0047 | 0.0024<br>SD=0.0016 | 0.0073<br>SD=0.0059 | 0.0024<br>SD=0.0016 | 0.0073<br>SD=0.0059 |
| Systemic lupus erythematosus | 0.0282<br>SD=0.0055 | 0.0682<br>SD=0.0245 | 0.0318<br>SD=0.0049 | 0.0709<br>SD=0.0220 | 0.0318<br>SD=0.0049 | 0.0709<br>SD=0.0220 |
| Primary biliary cholangitis | 0.0230<br>SD=0.0081 | 0.0668<br>SD=0.0397 | 0.0177<br>SD=0.0058 | 0.0370<br>SD=0.0408 | 0.0177<br>SD=0.0058 | 0.0370<br>SD=0.0408 |
| Vitiligo | 0.0379<br>SD=0.0156 | 0.1163<br>SD=0.0566 | N/A | N/A | N/A | N/A |
| Alopecia areata | 0.0061<br>SD=0.0038 | 0.0083<br>SD=0.0219 | 0.0053<br>SD=0.0037 | 0.0075<br>SD=0.0180 | 0.0052<br>SD=0.0037 | 0.0074<br>SD=0.0179 |
| Psoriasis | 0.0233<br>SD=0.0028 | 0.0736<br>SD=0.0117 | 0.0118<br>SD=0.0014 | 0.0350<br>SD=0.0103 | 0.0118<br>SD=0.0014 | 0.0350<br>SD=0.0103 |
| Adrenocortical insufficiency | 0.0166<br>SD=0.0049 | 0.0866<br>SD=0.0263 | 0.0098<br>SD=0.0040 | 0.0568<br>SD=0.0223 | 0.0098<br>SD=0.0040 | 0.0567<br>SD=0.0223 |
| IgA nephropathy | 0.0016<br>SD=0.0013 | -0.0123<br>SD=0.0124 | N/A | N/A | 0.0026<br>SD=0.0029 | -0.0091<br>SD=0.0145 |
| Idiopathic thrombocytopenic purpura | 0.0014<br>SD=0.0011 | -0.0024<br>SD=0.0100 | 0.0015<br>SD=0.0010 | -0.0038<br>SD=0.0102 | 0.0015<br>SD=0.0010 | -0.0038<br>SD=0.0104 |

| | | | | | | |
|---|---|---|---|---|---|---|
| Ankylosing spondylitis | 0.1241<br>SD=0.0064 | 0.2476<br>SD=0.0232 | 0.0951<br>SD=0.0060 | 0.2292<br>SD=0.0221 | 0.0951<br>SD=0.0060 | 0.2292<br>SD=0.0221 |
| Systemic sclerosis | 0.0021<br>SD=0.0012 | -0.0075<br>SD=0.0113 | 0.0020<br>SD=0.0022 | -0.0052<br>SD=0.0119 | 0.0020<br>SD=0.0022 | -0.0052<br>SD=0.0119 |
| Inflammatory bowel disease | 0.0071<br>SD=0.0010 | 0.0267<br>SD=0.0093 | 0.0041<br>SD=0.0008 | 0.0167<br>SD=0.0087 | 0.0043<br>SD=0.0009 | 0.0176<br>SD=0.0088 |
| Wegener granulomatosis | 0.0081<br>SD=0.0055 | 0.0184<br>SD=0.0193 | 0.0050<br>SD=0.0051 | 0.0021<br>SD=0.0171 | 0.0050<br>SD=0.0051 | 0.0021<br>SD=0.0173 |
| Myasthenia gravis | 0.0019<br>SD=0.0018 | -0.0127<br>SD=0.0170 | 0.0012<br>SD=0.0025 | -0.0091<br>SD=0.0143 | 0.0012<br>SD=0.0025 | -0.0090<br>SD=0.0146 |
| Allergic purpura | 0.0022<br>SD=0.0022 | 0.0065<br>SD=0.0148 | 0.0022<br>SD=0.0025 | 0.0048<br>SD=0.0171 | N/A | N/A |
| Sjögren syndrome | 0.0099<br>SD=0.0031 | 0.0208<br>SD=0.0103 | 0.0068<br>SD=0.0024 | 0.0140<br>SD=0.0090 | 0.0068<br>SD=0.0024 | 0.0140<br>SD=0.0090 |
| Guillain-Barre syndrome | 0.0019<br>SD=0.0012 | -0.0063<br>SD=0.0089 | 0.0018<br>SD=0.0012 | -0.0066<br>SD=0.0102 | 0.0018<br>SD=0.0012 | -0.0067<br>SD=0.0104 |
| Autoimmune hemolytic anemia | 0.0013<br>SD=0.0017 | -0.0057<br>SD=0.0077 | 0.0009<br>SD=0.0011 | -0.0047<br>SD=0.0053 | 0.0009<br>SD=0.0011 | -0.0047<br>SD=0.0053 |
| Multiple sclerosis | 0.0396<br>SD=0.0068 | 0.0897<br>SD=0.0213 | 0.0319<br>SD=0.0067 | 0.0761<br>SD=0.0222 | 0.0319<br>SD=0.0067 | 0.0761<br>SD=0.0222 |

##### 3.3.4 Analyses in non-HLA regions – LDSC

**Table S3.3.4.1: Genetic correlations between T1D and other 25 AIDs**

| Disease | Rg (95% CI) | P-value |
|---|---|---|
| Autoimmune hypothyroidism | 0.43 [0.34, 0.51] | 2.85E-23 |
| Coeliac disease | 0.27 [0.13, 0.42] | 1.93E-04 |
| Rheumatoid arthritis | 0.43 [0.25, 0.61] | 3.88E-06 |
| Vitamin B12 deficiency anaemia | 0.48 [0.34, 0.63] | 6.35E-11 |
| Sarcoidosis | 0.25 [0.12, 0.38] | 9.90E-05 |
| Autoimmune hyperthyroidism | 0.33 [0.18, 0.48] | 1.21E-05 |
| Mixed connective tissue disease | -0.18 [-0.36, -0.01] | 3.95E-02 |
| Sjögren syndrome | 0.17 [0.04, 0.30] | 9.20E-03 |
| Vitiligo | 0.31 [0.19, 0.42] | 2.56E-07 |
| Primary biliary cholangitis | 0.39 [0.22, 0.56] | 3.83E-06 |
| Autoimmune hemolytic anemia | -0.63 [-1.00, 1.00] | 8.16E-01 |
| Myasthenia gravis | 0.27 [0.10, 0.43] | 1.93E-03 |
| Adrenocortical insufficiency | -0.11 [-0.49, 0.27] | 5.69E-01 |
| Psoriasis | 0.11 [0.03, 0.19] | 1.05E-02 |
| Systemic lupus erythematosus | 0.33 [0.19, 0.47] | 5.03E-06 |
| IgA nephropathy | -0.08 [-0.25, 0.09] | 3.68E-01 |
| Systemic sclerosis | -0.35 [-0.77, 0.07] | 1.02E-01 |
| Alopecia areata | 0.32 [0.11, 0.54] | 2.70E-03 |
| Guillain-Barre syndrome | 0.14 [-0.12, 0.4] | 2.99E-01 |
| Allergic purpura | -0.23 [-0.64, 0.17] | 2.54E-01 |
| Inflammatory bowel disease | 0.00 [-0.02, 0.03] | 8.13E-01 |
| Wegener granulomatosis | -0.16 [-0.57, 0.26] | 4.55E-01 |
| Ankylosing spondylitis | 0.11 [-0.04, 0.25] | 1.45E-01 |
| Idiopathic thrombocytopenic purpura | 0.17 [-0.18, 0.51] | 3.37E-01 |
| Multiple sclerosis | 0.17 [-0.12, 0.46] | 2.57E-01 |

**Table S3.3.4.2: Sensitivity analysis of genetic correlations between T1D and other AIDs significantly correlated with T1D**

| Trait 1 | Trait 2 | Include chromosome 6 | | Exclude chromosome 6 | |
|---|---|---|---|---|---|
| | | Rg | p | Rg | p |
| Type 1 diabetes | Rheumatoid arthritis | 0.46 | 0.0010 | 0.47 | 0.0000 |
| Type 1 diabetes | Autoimmune hyperthyroidism | 0.48 | 0.0000 | 0.47 | 0.0000 |
| Type 1 diabetes | Autoimmune hypothyroidism | 0.60 | 0.0000 | 0.60 | 0.0000 |
| Type 1 diabetes | Coeliac disease | 0.27 | 0.0002 | 0.25 | 0.0005 |
| Type 1 diabetes | Vitamin B12 deficiency anaemia | 0.64 | 0.0000 | 0.62 | 0.0000 |
| Type 1 diabetes | Sjögren's syndrome | 0.39 | 0.0000 | 0.40 | 0.0000 |

##### 3.4 Inter-generation cross-trait transmission

##### Table S3.4.1: Associations between non-HLA PGS and AIDs at population level

| | Association between an AID and non-HLA polygenic risk for this AID | | |
|---|---|---|---|
| **Disease** | **OR (95% CI)** | **P-value** | **ρ(%)** |
| Type 1 diabetes | 2.29 [2.22, 2.37] | 1.00E-519 | 7.93 |
| Autoimmune hypothyroidism | 1.73 [1.71, 1.75] | 1.00E-2397 | 16.00 |
| Autoimmune hyperthyroidism | 1.25 [1.21, 1.3] | 7.89E-39 | 1.95 |
| Vitamin B12 deficiency anaemia | 1.24 [1.2, 1.29] | 4.15E-40 | 1.97 |
| Sarcoidosis | 1.27 [1.23, 1.3] | 4.59E-59 | 2.43 |
| Primary biliary cholangitis | 1.27 [1.18, 1.37] | 5.15E-10 | 0.93 |
| Coeliac disease | 1.66 [1.61, 1.72] | 9.73E-184 | 4.34 |
| Inflammatory bowel disease | 1.58 [1.55, 1.6] | 1.00E-582 | 7.78 |
| IgA nephropathy | 1.07 [1, 1.15] | 6.03E-02 | 0.28 |
| Ankylosing spondylitis | 1.58 [1.52, 1.64] | 2.76E-120 | 3.48 |
| Rheumatoid arthritis | 1.46 [1.43, 1.48] | 1.00E-426 | 6.64 |
| Sjögren syndrome | 1.18 [1.14, 1.23] | 1.18E-19 | 1.36 |
| Systemic sclerosis | 1.17 [1.07, 1.28] | 5.30E-06 | 0.70 |
| Systemic lupus erythematosus | 1.76 [1.66, 1.86] | 1.42E-83 | 2.91 |
| Multiple sclerosis | 1.79 [1.72, 1.86] | 1.91E-190 | 4.42 |
| Myasthenia gravis | 1.23 [1.13, 1.35] | 5.24E-06 | 0.68 |
| Alopecia areata | 1.05 [0.99, 1.13] | 1.19E-01 | 0.23 |
| Psoriasis | 1.5 [1.48, 1.53] | 1.00E-398 | 6.41 |
| Vitiligo | 1.54 [1.39, 1.72] | 5.82E-15 | 1.17 |

##### Table S3.4.2 Polygenic transmission disequilibrium tests by HLA and non-HLA in trios

| Disease | Region | Group status | Count | Mean (95% CI) | P-value |
|---|---|---|---|---|---|
| Type 1 diabetes | HLA | Affected | 779 | 1.23 [1.16, 1.3] | 2.61E-163 |
| Type 1 diabetes | HLA | Unaffected | 1321 | -0.06 [-0.13, 0.01] | 8.19E-02 |
| Type 1 diabetes | non-HLA | Affected | 779 | 0.69 [0.62, 0.75] | 9.59E-74 |
| Type 1 diabetes | non-HLA | Unaffected | 1321 | 0 [-0.06, 0.05] | 8.79E-01 |
| Autoimmune hypothyroidism | HLA | Affected | 565 | 0.4 [0.31, 0.49] | 5.74E-18 |
| Autoimmune hypothyroidism | HLA | Unaffected | 935 | 0 [-0.07, 0.07] | 9.62E-01 |
| Autoimmune hypothyroidism | non-HLA | Affected | 565 | 0.28 [0.2, 0.36] | 8.02E-11 |
| Autoimmune hypothyroidism | non-HLA | Unaffected | 935 | -0.04 [-0.11, 0.03] | 2.26E-01 |
| Coeliac disease | HLA | Affected | 768 | 0.67 [0.59, 0.74] | 1.74E-62 |
| Coeliac disease | HLA | Unaffected | 1269 | -0.02 [-0.08, 0.05] | 5.87E-01 |
| Coeliac disease | non-HLA | Affected | 768 | 0.04 [-0.02, 0.1] | 2.23E-01 |
| Coeliac disease | non-HLA | Unaffected | 1269 | 0.01 [-0.04, 0.05] | 4.42E-01 |

| | | | | | |
|---|---|---|---|---|---|
| Rheumatoid arthritis | HLA | Affected | 728 | 0.53 [0.46, 0.6] | 7.35E-43 |
| Rheumatoid arthritis | HLA | Unaffected | 1234 | -0.01 [-0.07, 0.06] | 8.44E-01 |
| Rheumatoid arthritis | non-HLA | Affected | 728 | 0.21 [0.14, 0.28] | 5.16E-09 |
| Rheumatoid arthritis | non-HLA | Unaffected | 1234 | -0.08 [-0.14, -0.02] | 6.00E-03 |
| Systemic lupus erythematosus | HLA | Affected | 797 | 0.27 [0.21, 0.34] | 3.08E-15 |
| Systemic lupus erythematosus | HLA | Unaffected | 1347 | 0.01 [-0.04, 0.07] | 6.54E-01 |
| Systemic lupus erythematosus | non-HLA | Affected | 797 | 0.13 [0.06, 0.19] | 1.58E-04 |
| Systemic lupus erythematosus | non-HLA | Unaffected | 1347 | -0.02 [-0.07, 0.02] | 3.24E-01 |
| Sarcoidosis | HLA | Affected | 781 | 0.12 [0.06, 0.19] | 3.23E-04 |
| Sarcoidosis | HLA | Unaffected | 1322 | -0.01 [-0.06, 0.04] | 7.12E-01 |
| Sarcoidosis | non-HLA | Affected | 781 | 0.11 [0.04, 0.18] | 1.41E-03 |
| Sarcoidosis | non-HLA | Unaffected | 1322 | 0 [-0.05, 0.05] | 9.59E-01 |
| Psoriasis | HLA | Affected | 751 | 0.13 [0.06, 0.2] | 3.91E-04 |
| Psoriasis | HLA | Unaffected | 1256 | -0.01 [-0.07, 0.04] | 6.54E-01 |
| Psoriasis | non-HLA | Affected | 751 | -0.01 [-0.07, 0.06] | 7.96E-01 |
| Psoriasis | non-HLA | Unaffected | 1256 | 0.03 [-0.02, 0.08] | 2.35E-01 |
| Inflammatory bowel disease | HLA | Affected | 756 | -0.38 [-0.45, -0.31] | 3.07E-22 |
| Inflammatory bowel disease | HLA | Unaffected | 1284 | -0.04 [-0.1, 0.02] | 1.74E-01 |
| Inflammatory bowel disease | non-HLA | Affected | 756 | -0.01 [-0.08, 0.06] | 8.05E-01 |
| Inflammatory bowel disease | non-HLA | Unaffected | 1284 | -0.01 [-0.07, 0.04] | 6.55E-01 |
| Multiple sclerosis | HLA | Affected | 793 | -0.15 [-0.23, -0.08] | 3.45E-05 |
| Multiple sclerosis | HLA | Unaffected | 1341 | 0.07 [0.02, 0.13] | 1.10E-02 |
| Multiple sclerosis | non-HLA | Affected | 793 | 0.05 [-0.02, 0.12] | 1.99E-01 |
| Multiple sclerosis | non-HLA | Unaffected | 1341 | -0.02 [-0.07, 0.04] | 5.97E-01 |
| Ankylosing spondylitis | HLA | Affected | 793 | -0.11 [-0.18, -0.04] | 1.27E-03 |
| Ankylosing spondylitis | HLA | Unaffected | 1337 | -0.02 [-0.08, 0.03] | 4.04E-01 |
| Ankylosing spondylitis | non-HLA | Affected | 793 | -0.09 [-0.16, -0.02] | 1.38E-02 |
| Ankylosing spondylitis | non-HLA | Unaffected | 1337 | -0.01 [-0.07, 0.05] | 7.23E-01 |
| Vitamin B12 deficiency anaemia | HLA | Affected | 778 | 0.39 [0.32, 0.45] | 2.65E-27 |
| Vitamin B12 deficiency anaemia | HLA | Unaffected | 1320 | 0.02 [-0.04, 0.07] | 5.42E-01 |
| Autoimmune hyperthyroidism | HLA | Affected | 783 | 0.14 [0.06, 0.21] | 2.14E-04 |
| Autoimmune hyperthyroidism | HLA | Unaffected | 1327 | 0.01 [-0.05, 0.07] | 6.97E-01 |
| Primary biliary cholangitis | HLA | Affected | 796 | 0.3 [0.23, 0.37] | 1.87E-16 |
| Primary biliary cholangitis | HLA | Unaffected | 1346 | 0 [-0.05, 0.06] | 9.18E-01 |
| Mixed connective tissue disease | HLA | Affected | 790 | 0.47 [0.41, 0.54] | 1.39E-38 |
| Mixed connective tissue disease | HLA | Unaffected | 1331 | 0.01 [-0.05, 0.07] | 8.05E-01 |
| Sjögren syndrome | HLA | Affected | 786 | 0.21 [0.14, 0.27] | 1.46E-09 |
| Sjögren syndrome | HLA | Unaffected | 1322 | 0.03 [-0.02, 0.09] | 2.72E-01 |
| Adrenocortical insufficiency | HLA | Affected | 795 | 0.62 [0.55, 0.68] | 9.11E-61 |
| Adrenocortical insufficiency | HLA | Unaffected | 1345 | -0.01 [-0.06, 0.04] | 6.73E-01 |

##### 3.5 Distribution ratio between HLA PGS and non-HLA PGS

After observing these two different patterns from pTDT (**Figure S3.5.1**), we then wonder if we can quantify the contribution ratio between HLA PGS and non-HLA PGS for cross-disease transmission by considering genetic architecture of target disease. We took the ten AIDs with both HLA PGS and non-HLA PGS in pTDT into account and removed psoriasis and sarcoidosis when we used the ten AID to predict T1D at individual level and set $|\rho|>1\%$ (**Table S3.5.2**). For the rest seven AIDs (T1D is not applicable), we found that we could use our equation to quantify the contribution ratio between HLA PGS and non-HLA PGS using $\rho_{\text{HLA PGS}}$ and $\rho_{\text{non-HLA PGS}}$ while considering the results from pTDT (**Table S3.5.3**). For instance, for RA from the first group, the ratio trained from real data is 0.71:0.29 and that from our equation is 0.76:0.24. For IBD from the group that non-HLA PGS is not transmitted, the ratios are 0.96:0.04 and 1.00:0.00. The results show that the ratios from $\boldsymbol{ratio}_{proposed}$ calculated from our equation are very close to the observed ratios from $\boldsymbol{ratio}_{imperical}$ (Pearson correlation $\boldsymbol{r}$=0.977). Our equation could provide a reliable ratio between HLA PGS and non-HLA PGS in the setting of cross-disease transmission.

##### Figure S3.5.1 Two different patterns observed from pTDT

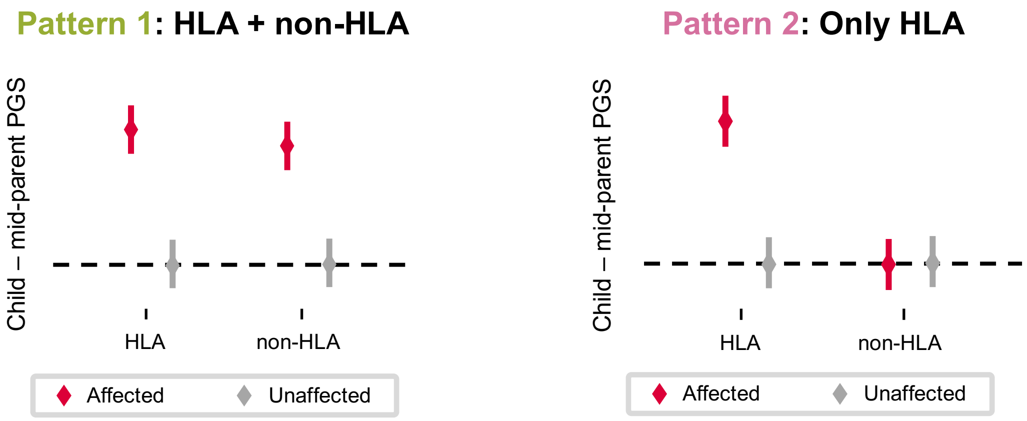


##### Table S3.5.2 Associations between PGSs for AIDs and T1D in population

| | HLA regions | | | non-HLA regions | | |
|---|---|---|---|---|---|---|
| **Disease** | **OR (95% CI)** | **P-value** | **ρ(%)** | **OR (95% CI)** | **P-value** | **ρ(%)** |
| T1D | 3.68 [3.54, 3.83] | 1.00E-959 | 12.54 | 1.84 [1.78, 1.91] | 1.12E-257 | 6.30 |
| HYPO | 1.55 [1.5, 1.6] | 2.86E-153 | 4.22 | 1.45 [1.4, 1.49] | 2.33E-104 | 3.40 |
| CD | 1.8 [1.75, 1.85] | 1.00E-421 | 7.61 | 1.1 [1.05, 1.14] | 5.11E-06 | 0.83 |
| RA | 1.89 [1.84, 1.95] | 1.00E-346 | 6.54 | 1.33 [1.29, 1.38] | 2.04E-63 | 2.66 |
| SLE | 1.3 [1.26, 1.34] | 1.02E-60 | 2.62 | 1.12 [1.08, 1.16] | 5.02E-11 | 1.10 |
| Sarcoidosis | 1.12 [1.08, 1.16] | 2.58E-11 | 0.97 | 1.09 [1.05, 1.13] | 3.60E-07 | 0.83 |
| Psoriasis | 1.11 [1.08, 1.15] | 8.15E-11 | 1.05 | 1.01 [0.97, 1.04] | 7.05E-01 | 0.01 |
| IBD | 0.65 [0.63, 0.67] | 4.53E-143 | -4.07 | 1.02 [0.98, 1.05] | 3.59E-01 | -0.13 |
| MS | 0.82 [0.79, 0.84] | 8.18E-30 | -1.77 | 1.02 [0.99, 1.06] | 2.33E-01 | 0.14 |
| AS | 0.97 [0.93, 1] | 6.00E-02 | -0.31 | 0.94 [0.91, 0.97] | 8.43E-05 | -0.69 |

#### Table S3.5.3 Contribution ratio between HLA PGS and non-HLA PGS

We reported Pearson correlation ($\boldsymbol{r}$) and three ratios between HLA PGS and non-HLA PGS. $\boldsymbol{ratio_{original}}$ is the ratio between $\rho_{\text{HLA PGS}}$ (the first $\rho$ in **Table S3.3.2.2**) and $\rho_{\text{non-HLA PGS}}$ ($\rho$ in **Table S3.4.1**) when we predicted AID using PGSs for that AID. $\boldsymbol{ratio_{imperical}}$ is from T1D prediction using the same PGSs ($\rho$ in **Table S3.5.2**). $\boldsymbol{ratio_{proposed}}$ is from our equation for T1D estimation (**Supplementary methods 2.5**).

| | $\boldsymbol{r}$ | $\boldsymbol{ratio_{original}}$ | | $\boldsymbol{ratio_{imperical}}$ | | $\boldsymbol{ratio_{proposed}}$ | |
|---|---|---|---|---|---|---|---|
| | | HLA | non-HLA | HLA | non-HLA | HLA | non-HLA |
| T1D | 0.128 | 0.67 | 0.33 | N/A | N/A | N/A | N/A |
| HYPO | 0.017 | 0.31 | 0.69 | 0.55 | 0.45 | 0.48 | 0.52 |
| RA | 0.079 | 0.61 | 0.39 | 0.71 | 0.29 | 0.76 | 0.24 |
| SLE | 0.104 | 0.48 | 0.52 | 0.70 | 0.30 | 0.65 | 0.35 |
| CD | 0.016 | 0.79 | 0.21 | 0.90 | 0.10 | 1.00 | 0.00 |
| Psoriasis | 0.038 | 0.55 | 0.45 | 0.99 | 0.01 | 1.00 | 0.00 |
| IBD | 0.005 | 0.29 | 0.71 | 0.96 | 0.04 | 1.00 | 0.00 |
| MS | 0.042 | 0.57 | 0.43 | 0.93 | 0.07 | 1.00 | 0.00 |

#### 3.6 Predicting risk of T1D in offspring using parental PGS

We tried five different AID PGSs to predict T1D at an individual level. Using trio data, we compared these five PGSs to clinical diagnoses from parents for T1D prediction in offspring. For each analysis, we reported their AUC, as well as McFadden's $R^2$, and the differences between PRS-CS and Full-PGS using P-value from t-test (P).

**Table S3.6.1 Comparisons among models using PRS-CS, Full-PGS using $ratio_{original}$, and HLA PGS along with non-HLA PGS at individual level for predicting the same AID**

| | | PRS-CS | | Full-PGS | | P | HLA + non-HLA | | HLA | | non-HLA | |
|---|---|---|---|---|---|---|---|---|---|---|---|---|
| | | AUC | R2 | AUC | R2 | | AUC | R2 | AUC | R2 | AUC | R2 |
| Pattern 1 | T1D | 0.804 | 0.187 | 0.906 | 0.364 | 3.02E-07 | 0.905 | 0.364 | 0.882 | 0.307 | 0.754 | 0.125 |
| | HYPO | 0.659 | 0.045 | 0.668 | 0.050 | 3.39E-01 | 0.668 | 0.050 | 0.568 | 0.008 | 0.655 | 0.043 |
| | RA | 0.613 | 0.019 | 0.672 | 0.045 | 1.74E-02 | 0.675 | 0.047 | 0.654 | 0.036 | 0.607 | 0.014 |
| | SLE | 0.617 | 0.022 | 0.649 | 0.024 | 5.50E-01 | 0.651 | 0.029 | 0.629 | 0.013 | 0.627 | 0.02 |
| Pattern 2 | CD | 0.578 | 0.009 | 0.831 | 0.140 | 4.40E-06 | 0.837 | 0.156 | 0.831 | 0.14 | 0.619 | 0.019 |
| | Psoriasis | 0.614 | 0.019 | 0.652 | 0.030 | 2.19E-01 | 0.666 | 0.036 | 0.628 | 0.02 | 0.618 | 0.018 |
| | IBD | 0.613 | 0.018 | 0.632 | 0.024 | 4.47E-01 | 0.646 | 0.029 | 0.605 | 0.015 | 0.599 | 0.014 |
| | MS | 0.597 | 0.016 | 0.669 | 0.033 | 1.11E-01 | 0.713 | 0.057 | 0.669 | 0.033 | 0.641 | 0.027 |

**Table S3.6.2 Comparisons among models using PRS-CS, Full-PGS using $ratio_{proposed}$, and HLA PGS along with non-HLA PGS at individual level for T1D prediction**

| | | PRS-CS | | Full-PGS | | P | HLA + non-HLA | | HLA | | non-HLA | |
|---|---|---|---|---|---|---|---|---|---|---|---|---|
| | | AUC | R2 | AUC | R2 | | AUC | R2 | AUC | R2 | AUC | R2 |
| Pattern 1 | T1D | 0.804 | 0.187 | 0.906 | 0.364 | 3.02E-07 | 0.905 | 0.364 | 0.882 | 0.307 | 0.754 | 0.125 |
| | HYPO | 0.631 | 0.032 | 0.668 | 0.051 | 5.29E-06 | 0.671 | 0.052 | 0.637 | 0.032 | 0.606 | 0.021 |
| | RA | 0.653 | 0.047 | 0.738 | 0.095 | 8.39E-04 | 0.738 | 0.095 | 0.73 | 0.085 | 0.592 | 0.016 |
| | SLE | 0.593 | 0.015 | 0.593 | 0.015 | 6.17E-01 | 0.593 | 0.015 | 0.578 | 0.012 | 0.551 | 0.005 |
| Pattern 2 | CD | 0.535 | 0.002 | 0.745 | 0.08 | 5.38E-08 | 0.745 | 0.08 | 0.745 | 0.08 | 0.513 | 0.000 |
| | Psoriasis | 0.511 | 0.000 | 0.553 | 0.002 | 6.21E-03 | 0.553 | 0.002 | 0.553 | 0.002 | 0.503 | 0.000 |
| | IBD | 0.545 | 0.004 | 0.639 | 0.032 | 6.85E-05 | 0.639 | 0.032 | 0.639 | 0.032 | 0.505 | 0.000 |
| | MS | 0.529 | 0.001 | 0.554 | 0.007 | 1.01E-01 | 0.554 | 0.007 | 0.554 | 0.007 | 0.503 | 0.000 |

**Table S3.6.3 AUC comparisons among models using parental diagnosis, parental full PGS using PRS-CS, Full-PGS, and parental HLA PGS along with parental non-HLA PGS in 12,563 genotyped trios for T1D prediction**

| | | Diagnosis | | PRS-CS | | Full-PGS | | P | HLA + non-HLA | | HLA | | non-HLA | |
|---|---|---|---|---|---|---|---|---|---|---|---|---|---|---|
| | | AUC | R2 | AUC | R2 | AUC | R2 | | AUC | R2 | AUC | R2 | AUC | R2 |
| Pattern 1 | T1D | 0.511 | 0.007 | 0.733 | 0.100 | 0.818 | 0.194 | 5.63E-05 | 0.817 | 0.194 | 0.793 | 0.159 | 0.684 | 0.063 |
| | HYPO | 0.527 | 0.002 | 0.59 | 0.016 | 0.613 | 0.024 | 6.31E-04 | 0.614 | 0.025 | 0.589 | 0.014 | 0.574 | 0.011 |
| | RA | 0.508 | 0.001 | 0.615 | 0.047 | 0.682 | 0.095 | 6.51E-03 | 0.682 | 0.095 | 0.674 | 0.085 | 0.572 | 0.016 |
| | SLE | 0.501 | 0.000 | 0.574 | 0.009 | 0.568 | 0.008 | 7.02E-01 | 0.566 | 0.008 | 0.546 | 0.003 | 0.546 | 0.003 |
| Pattern 2 | CD | 0.507 | 0.001 | 0.510 | 0.000 | 0.662 | 0.038 | 1.81E-06 | 0.662 | 0.038 | 0.662 | 0.038 | 0.505 | 0.000 |
| | Psoriasis | 0.508 | 0.001 | 0.509 | 0.000 | 0.530 | 0.001 | 5.07E-02 | 0.529 | 0.001 | 0.530 | 0.001 | 0.496 | 0.000 |
| | IBD | 0.503 | 0.000 | 0.522 | 0.001 | 0.592 | 0.015 | 2.91E-04 | 0.592 | 0.015 | 0.592 | 0.015 | 0.510 | 0.000 |
| | MS | 0.504 | 0.001 | 0.526 | 0.001 | 0.549 | 0.004 | 1.68E-01 | 0.550 | 0.004 | 0.549 | 0.004 | 0.509 | 0.000 |

**Figure S3.6.4 Mean Full-PGSs among affected children also overall had a larger deviation compared to mean PGS using PRS-CS**

For example, among T1D affected children, the mean full PGSs for CD using PRS-CS versus our approach deviated by 1.37 [1.31, 1.44] ($P=1.80\times10^{-188}$) and 0.87 [0.80, 0.94] ($P=3.00\times10^{-93}$).

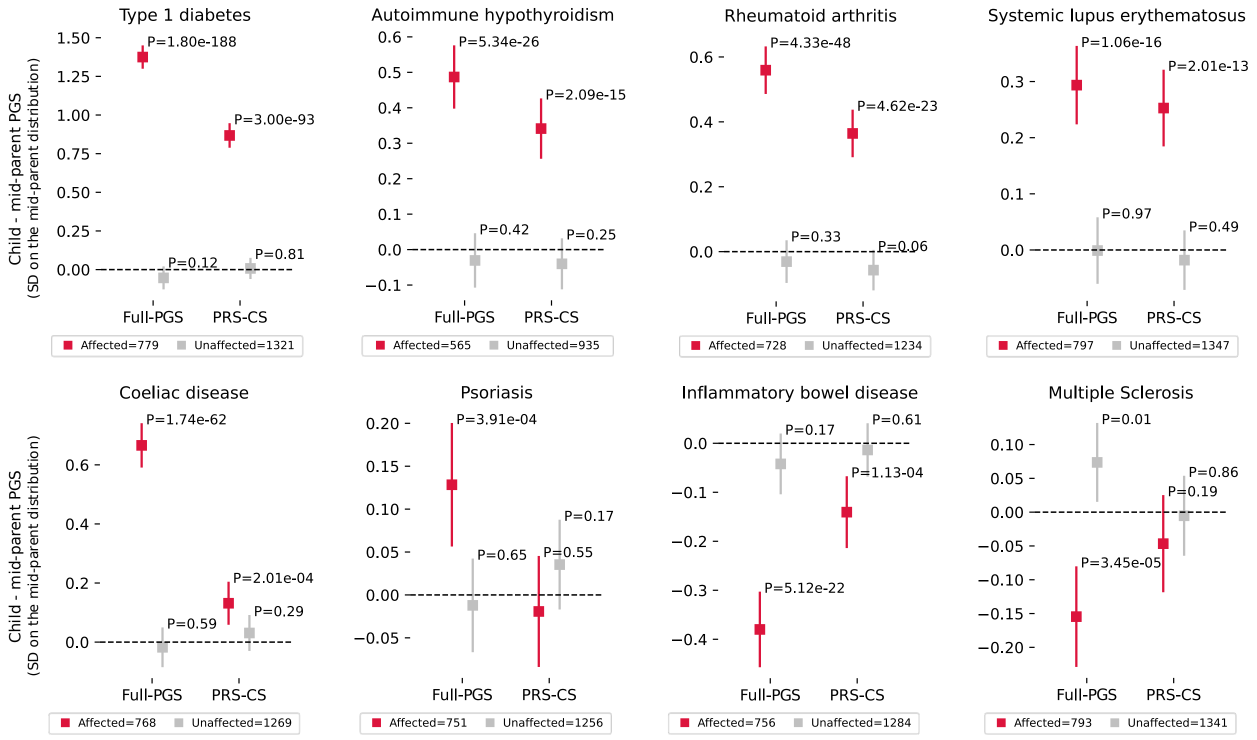

### 4. Supplementary discussion

**Table S4: Significant associations being reported from previous studies and in this study.**

we summarized our work and previous work reporting associations between familial aggregation of T1D and other AIDs using nationwide population-based studies with at least 2 000 AID cases (**Table S1**). Because of the different statistical methods and thresholds were applied in different studies, to ensure the statistics were in the same scaled framework, we defined a significant association according to P-value (positive association with P-value<0.05: *, positive association with P-value<0.01: **, negative association with P-value<0.01: --). From previous studies, an association can be either between parental AID and T1D in children or familial AID and T1D.

| Disease names | Previous nationwide registry-based studies | Registry-based study | | Genetic analyses | |
|---|---|---|---|---|---|
| | | T1D age<40 | T1D age<20 | HLA PGS | non-HLA $r_g$ |
| Type 1 diabetes | ** | ** | ** | ** | N/A |
| Autoimmune hypothyroidism | ** | ** | ** | ** | ** |
| Coeliac disease | ** | ** | ** | ** | ** |
| Rheumatoid arthritis | ** | ** | ** | ** | ** |
| Vitamin B12 deficiency anaemia | ** | ** | ** | | ** |
| Sarcoidosis | ** | ** | ** | ** | ** |
| Autoimmune hyperthyroidism | ** | ** | ** | ** | ** |
| Mixed connective tissue disease | | ** | ** | | * |
| Sjögren syndrome | ** | ** | | ** | ** |
| Vitiligo | | ** | * | | ** |
| Primary biliary cholangitis | ** | ** | ** | ** | ** |
| Autoimmune hemolytic anemia | | ** | | | |
| Myasthenia gravis | | * | | | ** |
| Adrenocortical insufficiency | ** | * | | ** | |
| Psoriasis | | * | | ** | * |
| Systemic lupus erythematosus | ** | | ** | ** | ** |
| IgA nephropathy | | | | | |

| | | |
|---|---|---|
| Systemic sclerosis | | |
| Alopecia areata | * | ** |
| Guillain-Barre syndrome | | |
| Allergic purpura | | |
| Inflammatory bowel disease ** | | -- |
| Wegener granulomatosis ** | | |
| Ankylosing spondylitis * | | |
| ITP | | |
| Multiple sclerosis ** | | -- |
